# The Genetic Architecture of Alzheimer’s Disease Risk: A Genomic Structural Equation Modelling Study

**DOI:** 10.1101/2021.02.23.21252211

**Authors:** Isabelle F Foote, Benjamin M Jacobs, Georgina Mathlin, Cameron J Watson, Phazha LK Bothongo, Sheena Waters, Ruth Dobson, Alastair J Noyce, Kamaldeep S Bhui, Ania Korszun, Charles R Marshall

## Abstract

**Background:** Targeting modifiable risk factors may have a role in the prevention of Alzheimer’s disease. However, the mechanisms by which these risk factors influence Alzheimer’s risk remain incompletely understood. Genomic structural equation modelling can reveal patterns of shared genetic architecture that provide insight into the pathophysiology of complex traits.

**Methods:** We identified genome-wide association studies for Alzheimer’s disease and its major modifiable risk factors: less education, hearing loss, hypertension, high alcohol intake, obesity, smoking, depression, social isolation, physical inactivity, type 2 diabetes, sleep disturbance and socioeconomic deprivation. We performed linkage disequilibrium score regression among these traits, followed by exploratory factor analysis, confirmatory factor analysis and structural equation modelling.

**Results:** We identified complex networks of linkage disequilibrium among Alzheimer’s disease risk factors. The data were best explained by a bi-factor model, incorporating a Common Factor for Alzheimer’s risk, and three orthogonal sub-clusters of risk factors, which were validated across the two halves of the autosome. The first sub-cluster was characterised by risk factors related to sedentary lifestyle behaviours, the second by traits associated with reduced life expectancy and the third by traits that are possible prodromes of Alzheimer’s disease. Alzheimer’s disease was more genetically distinct and displayed minimal shared genetic architecture with its risk factors, which was robust to the exclusion of *APOE*.

**Conclusion:** Shared genetic architecture may contribute to epidemiological associations between Alzheimer’s disease and its risk factors. Understanding the biology reflected by this communality may provide novel mechanistic insights that could help to prioritise targets for dementia prevention.

## Introduction

The rising prevalence of Alzheimer’s disease (AD) is a growing public health concern. However, estimates in several high-income countries show a decrease in age-specific incidence of dementia in recent birth cohorts [1-3]. This change has been attributed to improved access to education, reduction in the prevalence of cardiovascular disease and the uptake of healthy lifestyle behaviours, such as increased physical activity [1, 3, 4]. There is increasing interest in targeting dementia risk factors with a view to preventing or delaying the onset of dementia.

The recent Lancet Commission report on Dementia Prevention, Intervention and Care highlighted 12 potentially modifiable risk factors for dementia and estimated that elimination of these factors at key stages of life could prevent up to 40% of all-cause dementia cases [5]. However, the mechanisms by which these factors influence dementia risk, and whether they are truly causal, remain incompletely understood. Some factors such as sleep disturbance, social isolation, depression and hearing difficulty could, at least partly, serve as prodromal risk markers rather than causal risk factors [6-9]. An improved understanding of the causal pathways to clinical dementia development is urgently needed to support prevention efforts through the prioritisation of targets for intervention studies.

Advances in large, well-powered genome-wide association studies (GWASs) have shed light on the shared genetic architecture of complex traits and global pleiotropy. Significant bivariate genetic correlations measured by linkage disequilibrium (LD) score regression have been demonstrated between many behavioural and disease traits [10-14], indicating that extensive overlap in genetic pathways is present between seemingly distinct phenotypes. In the field of psychiatric genetics, genomic structural equation modelling (SEM) studies have identified patterns of shared genetic architecture between multiple psychiatric disorders, suggesting that these disorders have shared pathophysiological mechanisms driven by common pleiotropic risk variants in addition to disease-specific mechanisms [15, 16].

In this study, we aimed to apply these approaches to AD and its major risk factors within a multivariate model. We hypothesised there would be complex patterns of shared genetic architecture between AD and its risk factors, and that a factor analysis approach would suggest distinct aetiological pathways. This novel application could help to uncover shared or moderating risk factor pathways and will inform future work to better understand potential causal biological mechanisms that link modifiable risk factors to AD.

## Methods and Materials

### Trait selection and data formatting

In addition to Alzheimer’s disease itself, we selected AD risk factors that were identified by the Lancet Commission on Dementia Prevention, Intervention and Care [5]. The report identified 12 major modifiable risk factors, 10 of which were included in this analysis (less education, hearing loss, hypertension, high alcohol intake, obesity, smoking, depression, social isolation, physical inactivity and type 2 diabetes). Air pollution exposure was excluded as heritability estimates are low (all air pollution exposures had heritability estimates <0.015 in UK Biobank (https://nealelab.github.io/UKBB_ldsc/index.html)). We also excluded traumatic brain injury as there was not an appropriate GWAS available for this phenotype or a relevant proxy. We additionally included traits for sleep disturbance and socioeconomic deprivation. These were highlighted by the Lancet Commission as having strong associations with dementia risk, but with insufficient evidence for causality for them to be included in the final list of major modifiable risk factors [5]. We included two traits to measure social isolation (feelings of loneliness and less social activity) because existing literature indicates that these components of social isolation may have differential effects on dementia risk [7, 17].

GWAS summary statistics for each of the 14 included traits were identified (**Table 1** and **Supplementary Methods**) and formatted according to the pre-established requirements for LD score regression and genomic SEM [12, 15, 18]. To minimise bias from rare or poorly imputed alleles, we carried out uniform quality control according to the genomic SEM protocol [15] (**Supplementary Methods**). All summary statistics were derived from GWAS samples of unrelated individuals of European ancestry.

**Table 1:** Sample size and population prevalence for each of the included traits. The sample sizes may differ from the numbers reported in the original GWAS because we only used publicly available and European data. Population prevalence figures are current estimates for age-specific European ancestry populations (see **Supplementary Methods** for further details).

| Trait | Number of cases | Number of controls | Total sample size | Population prevalence | Original GWAS |
| --- | --- | --- | --- | --- | --- |
| Alzheimer's disease | 17,008 | 37,154 | 54,162 | 0.05 | Lambert et al [59] |
| Major depressive disorder | 59,851 | 113,154 | 173,005 | 0.13 | Wray et al [60] |
| Insomnia | 109,402 | 277,131 | 386,533 | 0.32 | Jansen et al [61] |
| Loneliness | 63,508 | 292,075 | 355,583 | 0.08 | http://www.nealelab.is/uk-biobank/ |
| Less social activity | 108,704 | 251,359 | 360,063 | 0.30 | http://www.nealelab.is/uk-biobank/ |
| Hearing difficulty | 134,141 | 219,842 | 353,983 | 0.39 | http://www.nealelab.is/uk-biobank/ |
| Less education | 61,093 | 296,456 | 357,549 | 0.27 | http://www.nealelab.is/uk-biobank/ |
| Physical inactivity | 21,255 | 338,008 | 359,263 | 0.18 | http://www.nealelab.is/uk-biobank/ |
| Smoking | 37,088 | 322,618 | 359,706 | 0.27 | http://www.nealelab.is/uk-biobank/ |
| Alcohol intake frequency | N/A | N/A | 360,726 | N/A | http://www.nealelab.is/uk-biobank/ |
| Body mass index | N/A | N/A | 354,831 | N/A | http://www.nealelab.is/uk-biobank/ |
| Deprivation status | N/A | N/A | 360,763 | N/A | http://www.nealelab.is/uk-biobank/ |
| Systolic blood pressure | N/A | N/A | 340,159 | N/A | http://www.nealelab.is/uk-biobank/ |
| Type 2 diabetes mellitus | 26,676 | 132,532 | 159,208 | 0.06 | Scott et al [62] |

### SNP-based heritability and genetic correlation estimation

We initially checked the univariate SNP-based heritability of each trait to ensure that they all had heritability Z scores >4 [12]. As heritability Z scores are influenced by the magnitude and precision of the heritability estimate, as well as GWAS sample size and the proportion of causal variants [19], using this value as a cut-off was preferable to basing inclusion on the heritability estimate alone. Heritability estimates were expressed on the liability scale for binary traits and the observed scale for continuous traits. Population prevalence for binary traits was defined based on recent estimates in European ancestry cohorts (**Table 1** and **Supplementary Methods**).

We then measured the genetic correlations between all pairs of traits to ensure that there was enough intercorrelation for genomic SEM to be appropriate, but that there were no correlations above 90% [20]. This was to ensure that we did not include highly multicollinear traits within our model, which may have induced bias towards a certain model specification due to near-complete genetic overlap between two traits [21]. We used a Bonferroni corrected significance threshold for 105 pairwise tests (*p* < 4.76E-4).

Both these steps were performed using *LDSC* software following pre-established protocols [12, 18] and calculated using publicly available LD scores and weights computed from 1000 Genomes European data restricted to Hapmap 3 SNPs with the major histocompatibility complex region removed.

### Genomic factor analysis

Genomic factor analysis was performed in R version 3.6.3. Because the underlying latent structure of genetic covariance between the included traits was unknown, we initially undertook a genomic exploratory factor analysis (EFA) to identify how many factors best explain the common genetic variance. We then used this to guide the specification of the parameters of a follow-up genomic confirmatory factor analysis (CFA).

Owing to the unavailability of suitable replication GWAS datasets for all of our traits, we ran the EFA in the odd autosomes and the CFA in the even autosomes as a cross-validation to minimise model overfitting. This approach has been utilised in a recent genomic SEM study [22]. It was felt to be appropriate in this context as all the traits included were polygenic, thus similar effects should be spread across the autosomes.

We undertook multivariable LD score regression to calculate the underlying genetic covariance matrix and its associated sampling covariance matrix in odd autosomal data to use as the input for EFA [15]. Using EFA we tested different numbers of factors to identify which model performed best using pre-defined criteria (**Supplementary Methods**). We used oblique (promax) rotation, which assumed correlation between factors, because the results of pairwise LD score regression indicated that there were high levels of genetic correlation between most traits, so it was unlikely that the latent constructs would be uncorrelated.

To test model fit we performed CFA using the output from an additional multivariable LD score regression in the even autosomes. We specified the parameters of the CFA using the results of the best performing model from EFA (i.e. the 3-factor solution). We examined multiple models within these parameters by comparing how inclusion of cross-loadings, negative loadings and higher loading cut-off levels influenced model fit. We assessed model fit by comparing recommended test results and cut-off points [15] (**Supplementary Methods**). From these results, we took the best performing model and using data from all autosomes we measured its overall genome-wide model fit.

We additionally tested the performance of a common factor model, a second-order model and a bi-factor model (**Supplementary Methods**). We ran the CFA and SEM models using diagonally weighted least squares (DWLS) estimation as this method accounts for differences in GWAS sample size and is more accurate for modelling binary traits [15, 23].

As AD susceptibility is highly influenced by the *APOE* gene, we also tested our model in all autosomes except chromosome 19 to assess how far the covariance in the model is driven by this area of the genome. Although SNPs with a χ^2^>30 are removed from LD score regression analysis [12], there is dense LD present between genes within this region so some *APOE*-related variance may still be captured via SNPs below this threshold [24, 25]. Therefore, we chose to take a conservative approach to measure non-*APOE*-related genetic architecture by excluding variants from chromosome 19 [26].

### Post-hoc sensitivity analyses

We noticed that the loadings for AD varied substantially between the EFA in odd autosomes, the CFA in even autosomes and the genome-wide SEM. We hypothesised that this may be because AD is more oligogenic than the other traits and so may show differential genetic correlations between chromosomes compared to highly polygenic traits which remain stable [26]. We therefore performed post-hoc sensitivity analyses to compare heritability and pairwise correlation estimates across individual chromosomes, compared EFA loading estimates between different chromosomal groupings and reran our models with AD excluded (**Supplementary Methods**).

## Results

### Heritability estimation and genetic correlations

All 14 traits displayed heritability Z scores >4, indicating that the traits were sufficiently heritable to be modelled using genomic SEM (**Table 2**).

**Table 2:** Results of the univariate LD heritability estimation for the 14 traits. SE = standard error.

| Trait | Observed or liability scale | Total SNP-based heritability (SE) | Heritability Z score | Mean $\chi^2$ | $\lambda$ GC | Intercept (SE) |
| --- | --- | --- | --- | --- | --- | --- |
| **Alzheimer's disease** | Liability | 0.0682 (0.0127) | 5.3701 | 1.1144 | 1.0926 | 1.0416 (0.0076) |
| **Major depressive disorder** | Liability | 0.0977 (0.006) | 16.2833 | 1.266 | 1.2365 | 0.9943 (0.009) |
| **Insomnia** | Liability | 0.084 (0.0038) | 22.1053 | 1.3659 | 1.3101 | 1.0152 (0.0089) |
| **Loneliness** | Liability | 0.0616 (0.0035) | 17.6 | 1.2766 | 1.2365 | 1.0162 (0.0074) |
| **Less social activity** | Liability | 0.0618 (0.0037) | 16.7027 | 1.2705 | 1.2365 | 1.0174 (0.0075) |
| **Hearing difficulty** | Liability | 0.0826 (0.0039) | 21.1795 | 1.3755 | 1.3068 | 1.024 (0.0085) |
| **Less education** | Liability | 0.2573 (0.0087) | 29.5747 | 1.7806 | 1.5995 | 1.0635 (0.0111) |
| **Physical inactivity** | Liability | 0.1292 (0.0095) | 13.6 | 1.1773 | 1.1619 | 1.0171 (0.007) |
| **Smoking** | Liability | 0.1969 (0.0102) | 19.3039 | 1.3734 | 1.3034 | 1.0142 (0.0084) |
| **Alcohol intake frequency** | Observed | 0.0797 (0.0036) | 22.1389 | 1.6203 | 1.4926 | 1.0521 (0.0105) |
| **Body mass index** | Observed | 0.2316 (0.0073) | 31.726 | 2.8005 | 2.1764 | 1.132 (0.0164) |
| **Deprivation status** | Observed | 0.0332 (0.0019) | 17.4737 | 1.2532 | 1.2299 | 1.0194 (0.0071) |
| **Systolic blood pressure** | Observed | 0.1403 (0.0052) | 26.9808 | 2.0666 | 1.6985 | 1.0909 (0.0134) |
| **Type 2 diabetes mellitus** | Liability | 0.121 (0.0084) | 14.4048 | 1.2311 | 1.1459 | 0.9979 (0.0087) |

We then measured SNP-based genetic correlations between all pairs of traits using LD score regression. Each trait shared at least one significant genetic correlation with another trait after Bonferroni correction, with most traits displaying positive correlations with multiple other traits, indicating a complex network of LD suitable for factor analysis (**Figure 1, Figure 2** and **Table S1**).

**Figure 1:**
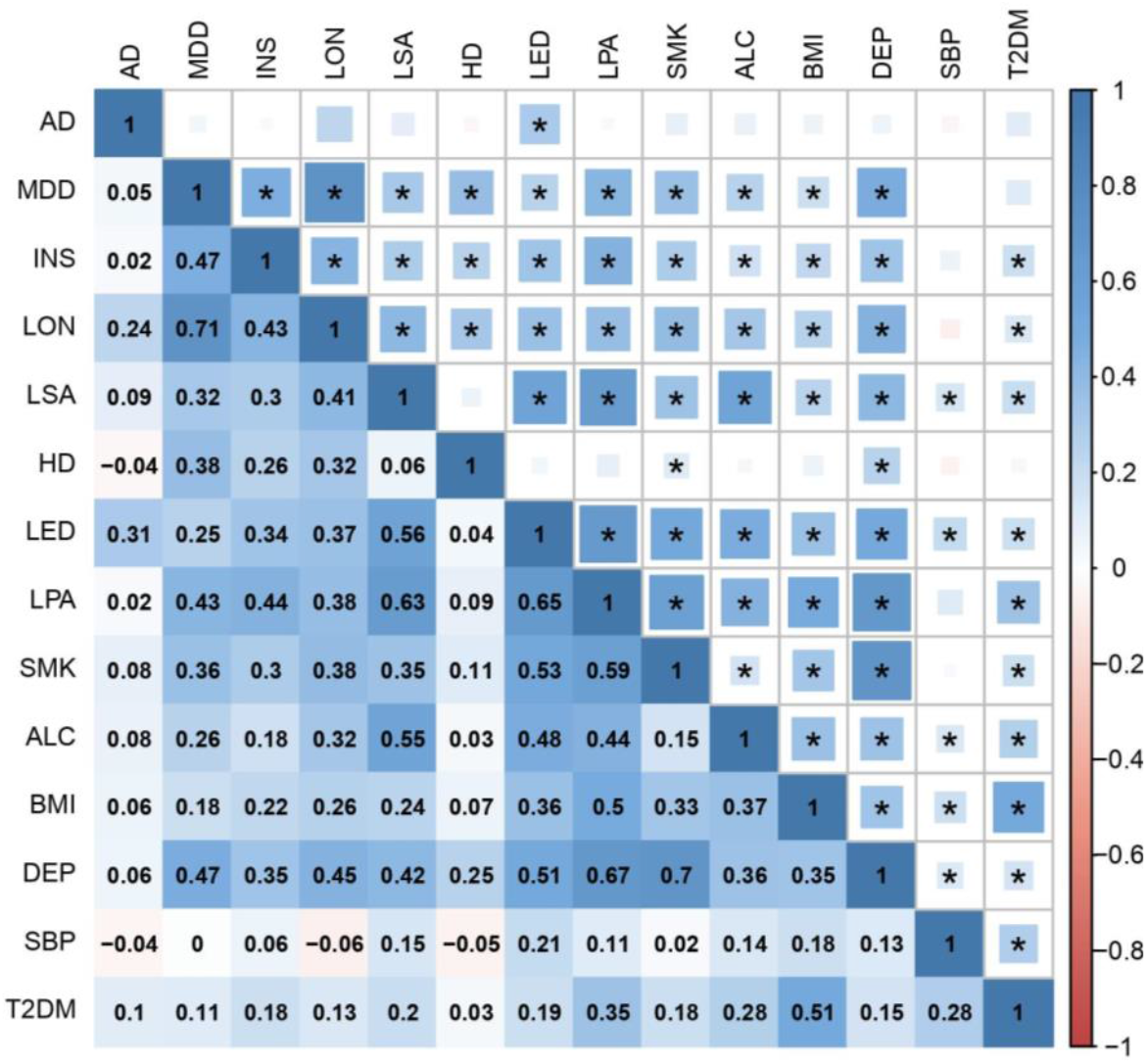
A heatmap of pairwise genome-wide genetic correlations between all 14 traits based on LD score regression. The upper triangle of the matrix displays the strength of correlation (size of each square) and significant associations passing Bonferroni correction (p < 4.76E-4) are marked with an asterisk. The lower triangle shows the correlation coefficient values. In both triangles, the shade of each square denotes a positive (blue) or negative (red) correlation, varying in shade by magnitude of correlation. AD Alzheimer’s disease; MDD major depressive disorder; INS insomnia; LON loneliness; LSA less social activity; HD hearing difficulty; LED less education; LPA physical inactivity; SMK smoking; ALC alcohol intake frequency; BMI body mass index; DEP deprivation status; SBP systolic blood pressure; T2DM type 2 diabetes mellitus.

**Figure 2:**
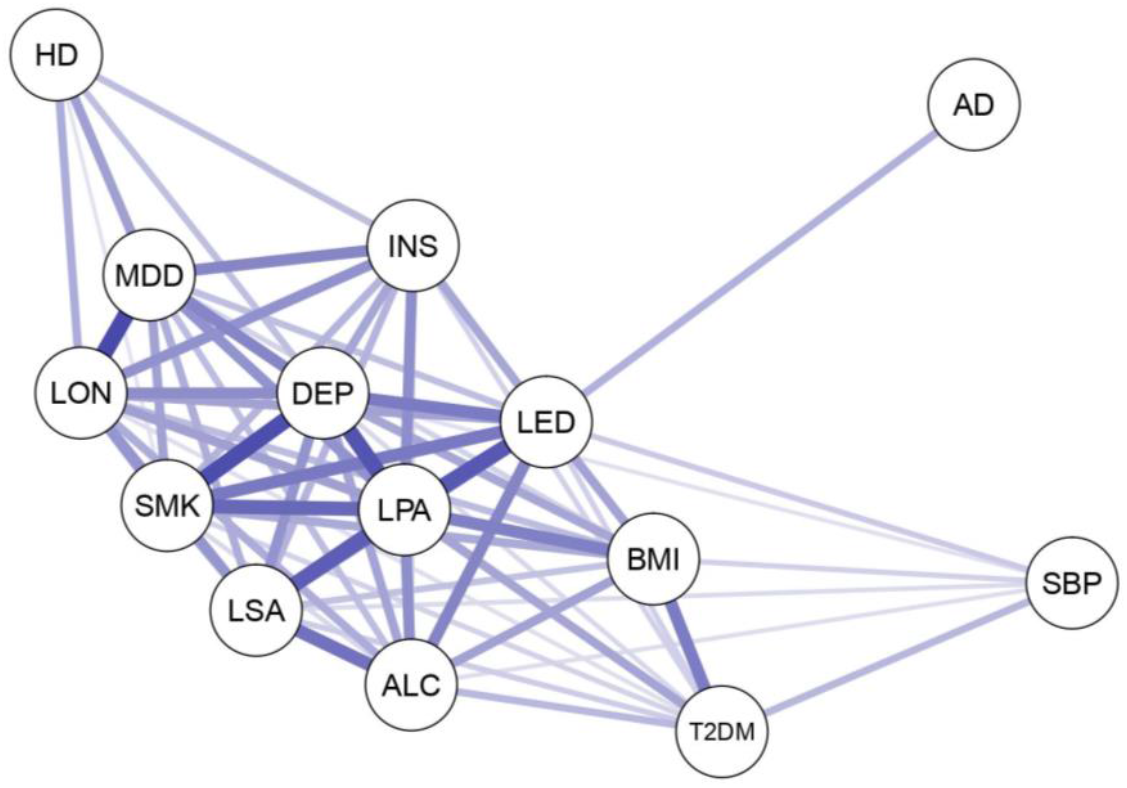
A weighted undirected graph showing the structure of the significant genetic correlations that are present between the 14 traits in our analysis. Each trait forms an individual node (depicted by a circle) and the lines joining them are edges which represent pairwise correlation coefficients between traits. Only Bonferroni significant relationships are included. The stronger the correlation, the shorter and wider the edge and the darker the colour. As all the significant correlations were positive, all edges are blue. AD Alzheimer’s disease; MDD major depressive disorder; INS insomnia; LON loneliness; LSA less social activity; HD hearing difficulty; LED less education; LPA physical inactivity; SMK smoking; ALC alcohol intake frequency; BMI body mass index; DEP deprivation status; SBP systolic blood pressure; T2DM type 2 diabetes mellitus.

### Factor analysis

EFA indicated that a 3-factor model fitted the data best (**Figure 3, Tables S2-S4** and **Supplementary Results**). Using the parameters of the 3-factor EFA, we ran CFA in the even chromosomes to establish model fit. As systolic blood pressure did not display any positive loadings above our pre-defined cut-off it was omitted from our subsequent models. The best fitting CFA included positive loadings ≥0.20, cross-loadings and between-factor correlations but excluded negative loadings (**Table 3**). Model fit improved when tested in all autosomes and after excluding chromosome 19 that contains *APOE* (**Figure 4a** and **Supplementary Results**).

**Table 3:** Model fit statistics for each of the genomic SEM models performed in even autosomes, all autosomes, all autosomes except chromosome 19 and all autosomes with Alzheimer’s disease (AD) excluded. None of these models included systolic blood pressure since it produced low loading estimates during exploratory factor analysis.

| **Autosomes used** | **Model type** | **EFA loading cut-off** | **Factor correlations (Y/N)** | **Cross loadings (Y/N)** | **Negative loadings (Y/N)** | **Degrees of freedom** | **X<sup>2</sup> statistic** | **Akaike Information Criterion (AIC)** | **Comparative Fit Index (CFI)** | **Standardised Root Mean Square Residual (SRMR)** |
| --- | --- | --- | --- | --- | --- | --- | --- | --- | --- | --- |
| Even | CFA | 0.2 | Y | Y | N | 54 | 485.6902 | 559.6902 | 0.8901 | 0.0620 |
| Even | CFA | 0.2 | Y | Y | Y | Model did not converge |  |  |  |  |
| Even | CFA | 0.2 | Y | N | Y | 74 | 992.9823 | 1054.982 | 0.7897 | 0.0838 |
| Even | CFA | 0.2 | Y | N | N | 62 | 606.4694 | 664.4694 | 0.8614 | 0.0736 |
| Even | CFA | 0.2 | N | Y | N | Model produced unstable model fit results due to negative residual variances |  |  |  |  |
| Even | CFA | 0.3 | Y | Y | N | Model produced unstable model fit results due to negative residual variances |  |  |  |  |
| Even | Common factor | NA | NA | NA | NA | 65 | 845.7252 | 897.7252 | 0.8013 | 0.0995 |
| Even | Second-order | 0.2 | NA | Y | N | 54 | 485.6897 | 559.6897 | 0.8901 | 0.0620 |
| Even | Bi-factor | 0.2 | NA | Y | N | 44 | 294.5661 | 388.5661 | 0.9362 | 0.0515 |
| All | CFA | 0.2 | Y | Y | N | 54 | 883.8453 | 957.8453 | 0.8839 | 0.0588 |
| All | Common factor | NA | NA | NA | NA | 65 | 1616.21 | 1668.21 | 0.7830 | 0.0975 |
| All | Second-order | 0.2 | NA | Y | N | Model did not converge |  |  |  |  |
| All | Bi-factor | 0.2 | NA | Y | N | 44 | 417.9627 | 511.9627 | 0.9477 | 0.0475 |
| All ex. 19 | CFA | 0.2 | Y | Y | N | 54 | 919.1961 | 993.1961 | 0.9039 | 0.0588 |
| All ex. 19 | Common factor | NA | NA | NA | NA | 65 | 1979.683 | 2031.683 | 0.7872 | 0.0979 |
| All ex. 19 | Second-order | 0.2 | NA | Y | N | 54 | 919.1951 | 993.1951 | 0.9039 | 0.0588 |
| All ex. 19 | Bi-factor | 0.2 | NA | Y | N | 44 | 479.8912 | 573.8912 | 0.9516 | 0.0473 |
| All (no AD) | CFA | 0.2 | Y | Y | N | 49 | 958.7412 | 1016.741 | 0.8604 | 0.0640 |
| All (no AD) | Common factor | 0.2 | NA | NA | N | 54 | 1512.458 | 1560.458 | 0.7762 | 0.1010 |
| All (no AD) | Second-order | 0.2 | NA | Y | N | 49 | 958.7427 | 1016.743 | 0.8604 | 0.0640 |
| All (no AD) | Bi-factor | 0.2 | NA | Y | N | 40 | 386.2076 | 462.2076 | 0.9469 | 0.0515 |

**Figure 3:**
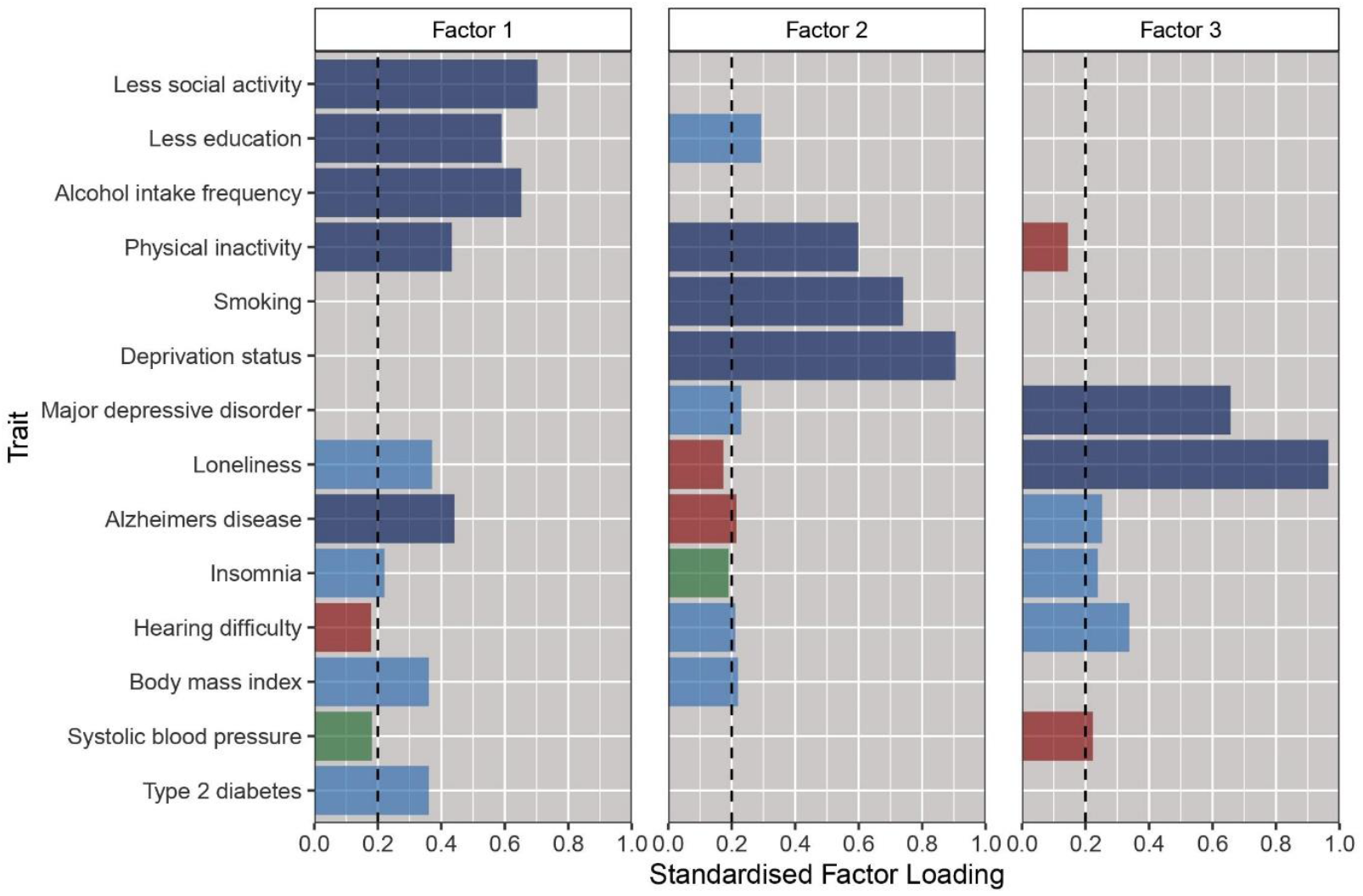
Standardised factor loadings for the 3-factor genomic exploratory factor analysis model based on data from the odd autosomes. Each bar on the plot denotes the factor loading of each trait on each of the 3 factors. Each vertical rectangle represents a factor. These factors are ordered by the amount of total variance they explain (left to right). The vertical dashed line represents the chosen cut-off threshold (0.20). The colour of the bar represents the factor loading strength. Dark blue denotes highly stable positive loadings (>0.40), light blue represents positive loadings between 0.20 to 0.40, green denotes positive loadings not meeting our cut-off threshold (between 0 and 0.20) and red represents negative loadings.

**Figure 4:**
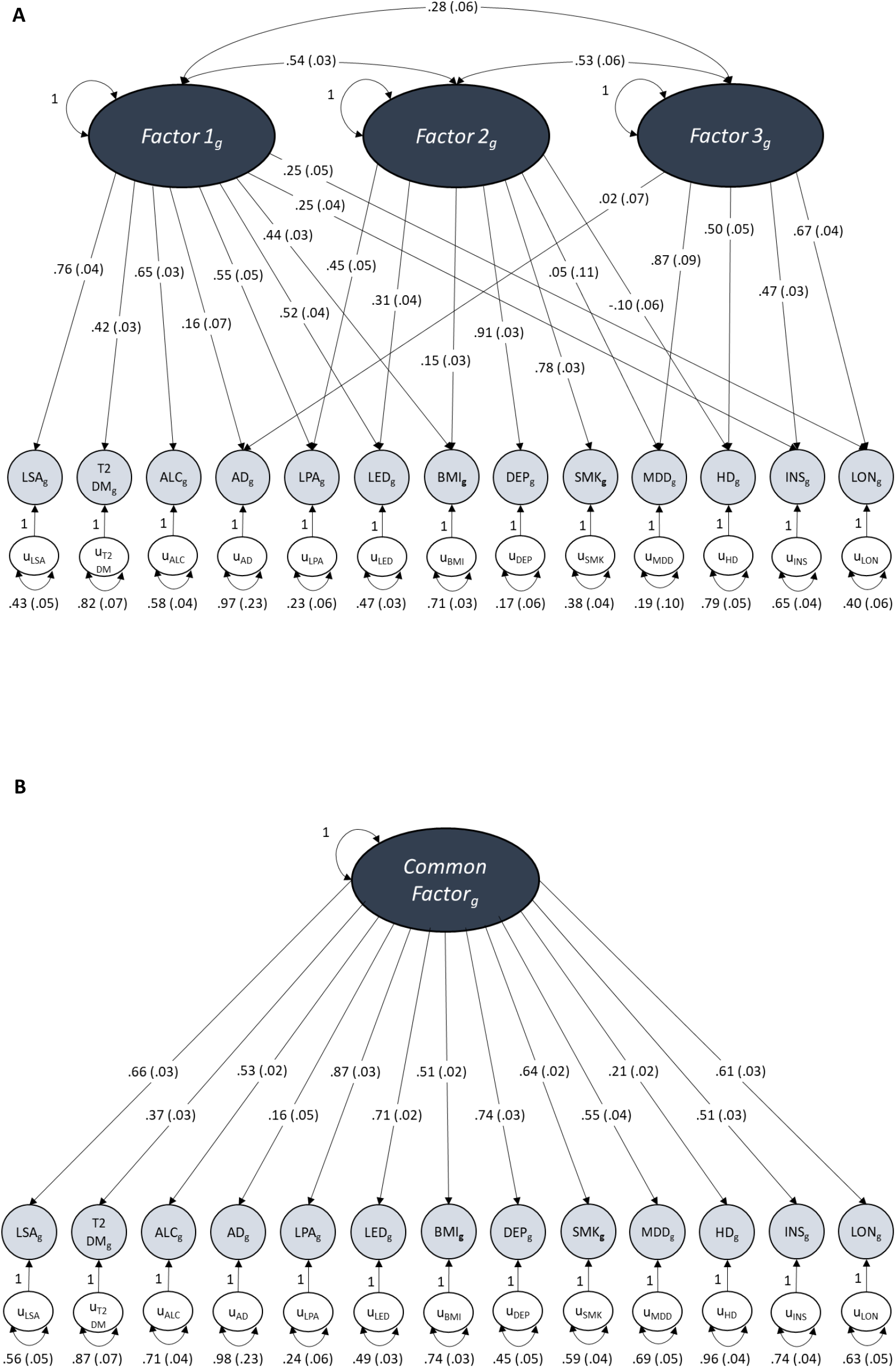
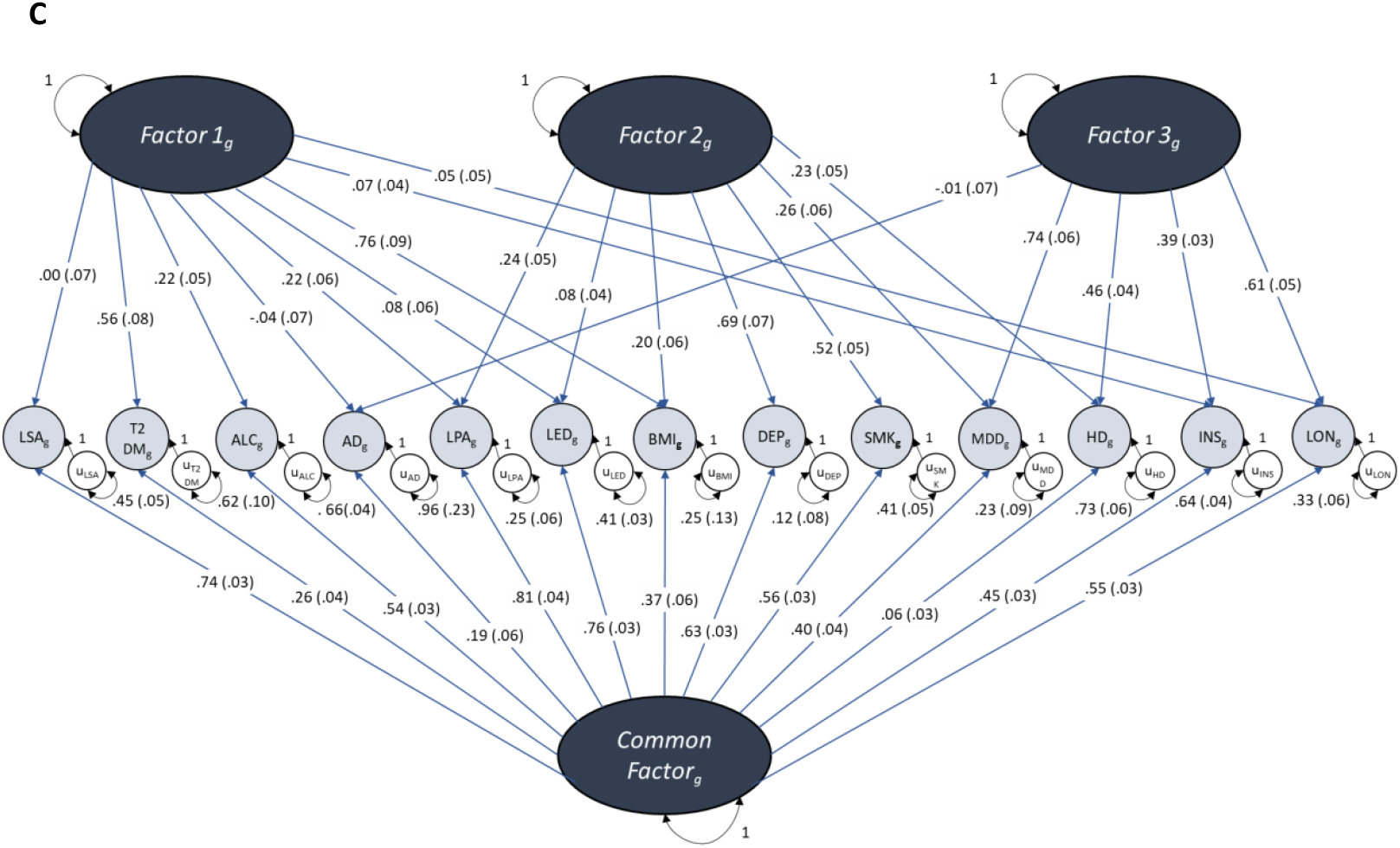
Path diagrams of the standardised solutions for the (a) 3-factor CFA, (b) common factor model and (c) bi-factor model conducted across all autosomes. The dark grey ovals represent latent unobserved constructs (factors), the pale grey circles are the observed traits made up of the measured genetic variance of each included trait, the white circles represent the unique variance of each trait not explained by loadings into factors. Unidirectional arrows depict regression coefficients from the independent variable to the dependent variable, curved two-headed arrows between factors represent correlations between the factors and two-headed arrows connecting a variable to itself highlights the variance of a variable with itself. Numbers in brackets represent the standard error of each measured value. Due to its lack of positive loading onto any factor during EFA, systolic blood pressure was excluded from these models. **Figure A** displays the path diagram for the 3-factor CFA which shows the factor loadings for traits that displayed a loading ≥ .20 at EFA. These three latent factors show moderate levels of inter-factor correlation. **Figure B** displays the path diagram for the common factor model where there is only one factor that depicts the overarching common variance between all included traits. **Figure C** displays the path diagram for the bi-factor model where there is a Common Factor of shared variance between all of the traits as well as 3 uncorrelated latent factors of sub-clusters of covariance that were specified by EFA in the same way as our CFA. AD Alzheimer’s disease; MDD major depressive disorder; INS insomnia; LON loneliness; LSA less social activity; HD hearing difficulty; LED less education; LPA physical inactivity; SMK smoking; ALC alcohol intake frequency; BMI body mass index; DEP deprivation status; T2DM type 2 diabetes mellitus.

Comparison of common factor, second-order factor and bi-factor models revealed that the bi-factor model provided the best model fit both in the even autosomes and across all autosomes (**Table 3, Figure 4** and **Figures S1-S2**). In the genome-wide bi-factor model, the Common Factor was influenced by all of the included traits except for hearing difficulty and the factors from the CFA formed orthogonal sub-clusters of variance between specific sets of AD risk factors that were independent of AD (**Figure 4c**). The model fit remained good after excluding chromosome 19 (**Table 3** and **Supplementary Results**).

### Post-hoc sensitivity analyses

Our sensitivity analyses found that in comparison to the risk factor traits, AD displayed substantial differences in SNP-based heritability estimates, pairwise genetic correlations with its risk factors and EFA loading estimates between the odd and the even chromosomes (**Tables S5-S6** and **Supplementary Results**). However, follow-up genomic factor analysis of data from all autosomes where AD and systolic blood pressure were excluded produced similar results to our main model, indicating that basing our model on data from the odd autosomes was appropriate for the AD risk factors and that the latent constructs are sufficiently stable to draw conclusions from (**Table S7** and **Supplementary Results**).

## Discussion

In this study, we modelled the shared genetic architecture between AD and its potentially modifiable risk factors. LD score regression highlighted pervasive genetic overlap between traits, though AD was more genetically unique than its risk factors. We built on this by using factor analysis and genomic SEM to model the pattern of this shared genetic architecture. The best fitting bi-factor model demonstrated the presence of a common genetic liability between AD and its risk factors, while the remaining genetic covariance was explained by three orthogonal clusters of traits. The findings demonstrate a richly interconnected network of genetic communality which may, in part, explain established epidemiological associations and suggest new avenues for both understanding the mechanisms by which modifiable risk factors influence AD, and aid the discovery of the components of missing heritability.

Pairwise bivariate LD score regression revealed many significant genetic correlations between the traits, indicating a substantial shared genetic component between AD risk factors. However, Alzheimer’s disease, systolic blood pressure and hearing difficulty had more limited and weaker genetic correlation with the other traits, suggesting that they are weaker genetic instruments and/or are more distinct in their genetic architecture. Our findings here support previous work that found AD to have a greater amount of unique genetic variance compared to psychiatric, behavioural or cardiometabolic traits [11, 12]. The strongest bivariate correlation for AD was with less education, which is consistent with previous Mendelian randomisation results, suggesting a causal effect of less education on risk of AD [27-29]. Our study goes beyond such bivariate approaches by identifying clusters of shared variance between traits that might not meet the stringent power requirements of Mendelian randomisation [30, 31], and highlighting smaller, yet potentially meaningful, overlap in genetic pathways.

Factor analysis identified an optimal 3-factor model, in which each factor contained a distinct cluster of traits, allowing us to generate some hypotheses about the mechanisms that might be reflected in these factors. Factor 1 had positive loadings for AD and a majority of the risk factors, suggesting that it might represent a common genetic pathway to AD across metabolic, psychiatric and lifestyle traits. Factor 2 had highly stable positive loadings for risk factors that are strongly associated with reduced life expectancy (smoking, deprivation and physical inactivity) [32-34] together with negative loading for AD. This factor might reflect only an apparent protective effect of these factors on AD due to premature mortality and survival bias [35-37], or could simply reflect a clustering of these traits independent of AD. Factor 3 included positive loadings for AD and a subset of risk factors that have complex relationships with AD in that they have all been hypothesised to be disease prodromes as well as causal risk factors [6-9, 38]. Moreover, systolic blood pressure negatively loaded onto this factor, and whereas risk of dementia is associated with higher blood pressure in mid-life, blood pressure declines during the period prior to cognitive symptoms and dementia diagnosis [39-42]. It is therefore possible that Factor 3 reflects AD prodromes, and hence reverse causality between AD and these traits.

A bi-factor model provided the best model fit in SEM. This model highlighted a Common Factor of shared genetic liability between the included traits in addition to the 3 distinct orthogonal clusters of covariance identified in the factor analysis. The only trait that did not pass inclusion into this model was systolic blood pressure due to its lack of communality with the other traits. This may partly be due to the complex relationship with blood pressure at the different stages of AD [39-42]. AD loaded positively onto the Common Factor but did not display positive loadings for any of the 3 sub-factors after the Common Factor was added to the model. This indicates that there is a common genetic liability between AD and its risk factors, but there is also a high degree of genetic overlap between different sets of AD risk factors independent of AD genetic pathways. The Common Factor had particularly high associations with being less educated, less physically active, more deprived and more socially isolated, indicating that these risk factors are those which have the highest degree of shared genetic liability with AD. However, the level of directly shared genetic architecture between risk factors and AD is likely to be low since 96% of the variance associated with AD was unique. The only risk factor that did not display a substantial loading onto the Common Factor was hearing difficulty. This may reflect that this risk factor has a limited role in determining AD risk, but rather is associated by reverse causation [43, 44].

The three orthogonal clusters identified in the factor analysis were largely unchanged in the SEM model, except that the AD loadings were attenuated because the shared variance between AD and its risk factors became entirely accounted for by the Common Factor. Additionally, Factor 1 only retained high loadings with traits associated with sedentary lifestyle behaviours (type 2 diabetes, higher alcohol intake, physical inactivity and obesity).

Despite their lack of direct shared genetic liability with AD, the likely relevance of these 3 sub-clusters is two-fold. Firstly, multimorbidity has been shown to increase an individual’s risk of developing AD, although the mechanisms for this remain unknown [45]. The high levels of shared genetic architecture between sets of known AD risk factors may represent mutual genetic liabilities between groups of traits that increase an individual’s likelihood of developing multiple conditions, or they might be associated with more generic ‘unhealthy’ behaviour and social circumstance. Secondly, although these clusters display no direct shared genetic pathway with AD, these shared pathways may exert pathophysiological effects that create a microenvironment within the brain that leads to an increased propensity to develop AD indirectly. In cases where an individual carries risk variants from multiple latent clusters, this may exert synergistic effects on AD development via non-genetic pathways, such as reducing cognitive reserve or via gene-environment interactions.

While the clustering of traits here may reflect causal genetic pathways between AD and certain risk factors, this analysis does not allow us to fully disentangle the nature of the observed pleiotropy, as we are unable to distinguish between causal, mediating and shared pathways. For example, since having less education is not only significantly genetically correlated with AD, but also all other risk factors (except for hearing difficulty), and it is one of the highest loading traits for the Common Factor, it seems probable that the common liability between less education may be a mediating or moderating driver of the shared relationship with AD and its other risk factors via extensive shared pleiotropy. Previous studies measuring bivariate associations have highlighted a causal genetic link between less education and AD [27-29] and its early life occurrence makes it a stronger candidate for being on the causal pathway to AD than other proposed risk factors that seem to exert risk later in life so are more likely to be linked by reverse causation [5]. Loneliness may also have a direct shared component with AD, but further work to identify the underlying SNPs in this model are needed to establish this. However, its high loading onto the Common Factor could moderate some of the shared liability seen for depression in this factor. Previous studies of the direct shared genetic architecture between depression and AD have been inconclusive [11, 46-48], so it is possible that loading of depression onto this factor is via its strong shared component with loneliness rather than by AD itself. Therefore, although we provide preliminary evidence of a potential shared liability between AD and risk factors, analysis of the SNP-based effects within this construct will enable a better understanding of how these traits are truly related on a genetic level and whether this is due to causal effects or shared risk variants.

There are several other important limitations to this work. Chief among these are the potential limitations of the genetic instruments, especially for AD. There was a high fluctuation of AD loading estimates between the CFA conducted in the even autosomes compared to the genome-wide estimates. Our sensitivity analyses indicated that this was driven by substantially different heritability and genetic correlation estimates between individual chromosomes. This may be due to the relatively oligogenic nature of AD compared to its risk factors [26], so shared genetic architecture may be confined to specific regions of the genome. This oligogenicity may not be appropriately modelled by methods designed for polygenic trait analysis, such as LD score regression, which expect risk variants to be spread evenly across the genome. Since LD score regression omits SNPs with a χ^2^>30, this could make AD a weaker genetic instrument by down-weighting the most important SNPs. This should be taken into consideration for genomic SEM analysis since the method bases its estimates on LD score regression and for certain traits, such as AD, it may not be suitable to split the chromosomes for factor analysis. However, post-hoc factor analysis of the more stable risk factor traits showed that the three latent constructs of shared genetic architecture between AD risk factors themselves are robust, so our conclusions relating to these constructs remain valid.

Furthermore, AD had the lowest heritability Z score of any of the traits that we measured. The precision of genetic correlations measured using LD score regression is influenced by sample size, whereas the magnitude of the correlation is not [12, 16], so the small sample size for AD probably contributed to the low Z score estimate. Heritability may also be adversely influenced by the heterogeneous nature of AD outcomes in the original GWAS [49]. The use of endophenotypes has been shown to produce more homogenous genetic phenotypes. However the current GWAS data for molecular biomarkers of AD pathology have poor heritability estimates and sample sizes too small to be used reliably for genomic SEM [50].

Another important limitation is that we have only used samples from European ancestry within our current analysis owing to the confounding effects of ancestral variation in LD score regression. Although sampling efforts in more diverse ancestral groups have started to increase, there is still a significant lack of data for many phenotypes for non-European groups and we were thus unable to perform an analysis that would include all our traits of interest [51]. Therefore, it is possible that these findings cannot be generalised to other populations. There is increasing evidence that AD risk factors exert differential effects on AD onset across diverse ethnicities [52, 53] and so, as sufficient data become available, it will be crucial to perform similar analyses in a range of ethnic groups to support targeted prevention strategies globally.

Lastly, our current model only measures shared genetic architecture, but does not identify shared environmental pathways or gene-environment interactions. There is evidence to suggest that an individual’s genetic profile can influence the magnitude of the effect that modifiable risk factors have on AD risk [54, 55]. Furthermore, increasing age remains the largest risk factor for AD and various epigenetic changes have been associated with both AD and ageing [56-58], so gene-environment interactions may be especially important to explore. However, such studies require a good understanding of the underlying genetic architecture, so our current study provides a useful and novel foundation for future work in this area.

This work provides the basis for further study to identify the SNPs and downstream pathways that are shared within the latent constructs identified here and those that are unique to AD and specific risk factors. GWAS of the factors within the model here may also yield new genetic associations with AD risk. Understanding these biological pathways that link AD and its risk factors could help to prioritise interventions that might prevent AD. The methods used here are also transferable to other disorders and risk factors so may provide further insight into a variety of diseases. Genomic SEM has proven successful in the field of psychiatric and behavioural genetics but has not been as widely adopted within other health research specialities and we hope this study will help show its potential merits. More broadly, future work in AD genetics should focus on phenotype optimisation and larger sample sizes of clinical AD cases that may lead to improved GWAS discovery and more links to risk factors may be unearthed. Larger GWASs of AD biomarkers, such as CSF amyloid-β and tau, will enable exploration of how AD risk factors share genetics with these specific pathologies, providing a deeper understanding of whether risk factors influence AD genetically via shared architecture with pathological processes or by reducing cognitive reserve. Lastly, analytical methods particularly suited to measuring oligogenic conditions will help to find the optimal way of modelling a relatively oligogenic disorder alongside highly polygenic ones.

Taken together, these results demonstrate a complex pattern of shared genetic architecture between AD and its risk factors, including a Common Factor for AD risk that may link pathophysiological mechanisms across metabolic, psychiatric and lifestyle traits. Understanding the biology underpinning this extensive communality could yield novel preventive strategies to mitigate the growing global challenge of dementia.

## Key Resources Table

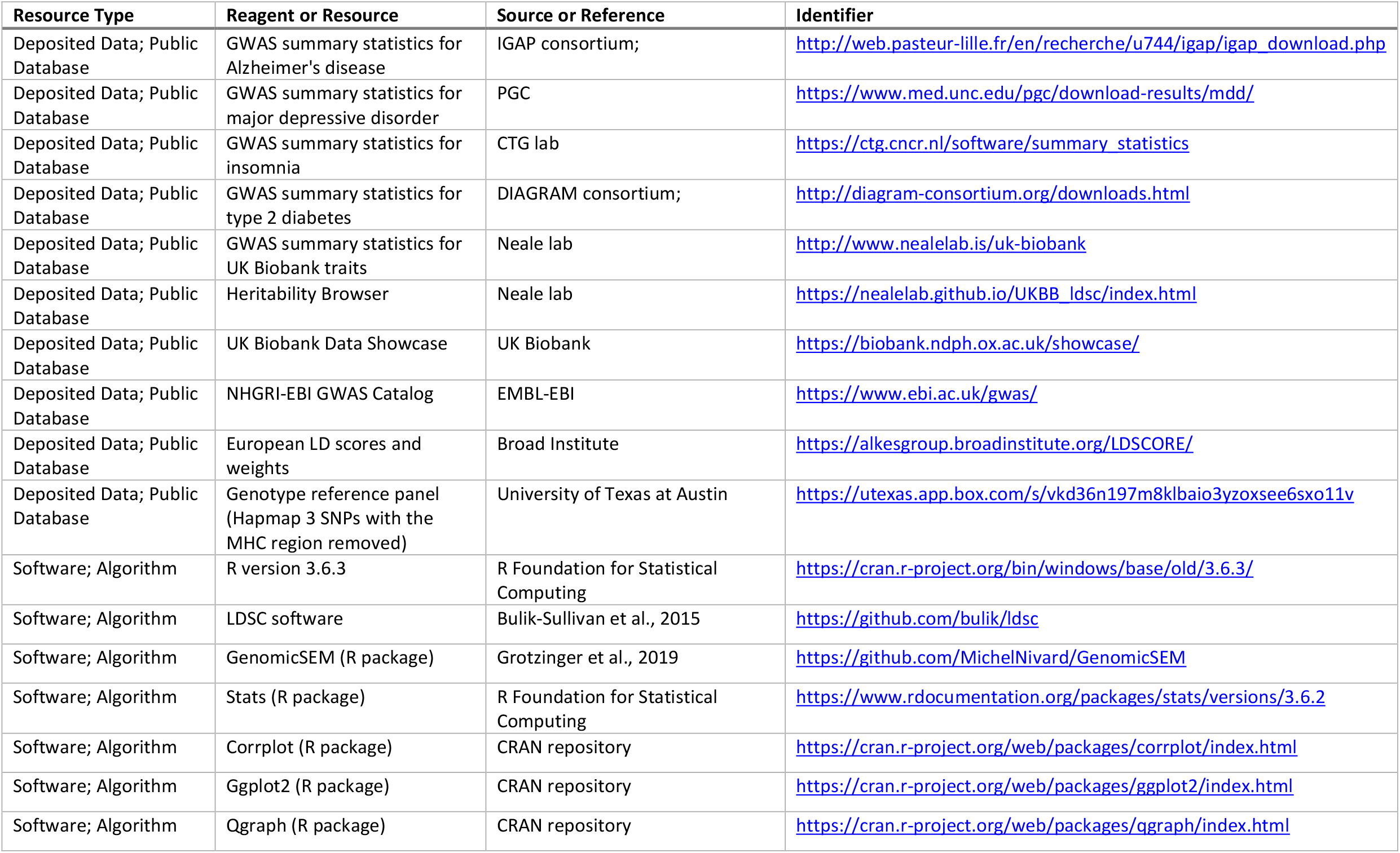

## Data Availability

The data used in the study are all publicly available for download (GWAS summary statistics, LD scores and weights, and the reference genome file). See the Key Resources Table in the main manuscript for details.

http://web.pasteur-lille.fr/en/recherche/u744/igap/igap_download.php

https://www.med.unc.edu/pgc/download-results/mdd/

https://ctg.cncr.nl/software/summary_statistics

http://diagram-consortium.org/downloads.html

http://www.nealelab.is/uk-biobank

https://alkesgroup.broadinstitute.org/LDSCORE/

https://utexas.app.box.com/s/vkd36n197m8klbaio3yzoxsee6sxo11v

## Acknowledgements

We would like to thank the participants in the included data cohorts that we used in this study and the researchers who conducted the original GWAS studies for making their data publicly accessible, thereby making our work possible. We also thank the developers of all the software we used for making this open source and special thanks to Andrew Grotzinger (University of Texas at Austin) for providing valuable technical support with the *GenomicSEM* software. We are grateful to the late Dr Robert Keers for his insightful comments on the analysis plan. This project was supported by the George Henry Woolfe Legacy Fund (WOL1000B) and a Bart’s Charity grant (MGU0366).

## Author contributions

IFF, CRM, AK and KSB created the study concept and design. IFF and CRM drafted the manuscript. IFF performed the data curation and analysis. AJN and BMJ provided technical advice on data analysis and genetic methods. GM guided aspects relating to factor analysis. RD provided critique and refinement on the methods. CJW, PLKB and SW provided input and theoretical knowledge with interpretation of results. All authors contributed to the final version of this manuscript. CRM, AK and KSB supervised this project.

