## Supplementary Materials for "The Genetic Architecture of Alzheimer’s Disease Risk: A Genomic Structural Equation Modelling Study"

#### Supplementary Methods

##### GWAS summary statistics identification and quality control procedures

GWAS summary statistics for each of the included traits were identified using GWAS Catalog (https://www.ebi.ac.uk/gwas/) and the UK Biobank Data Showcase (https://biobank.ndph.ox.ac.uk/showcase/) and formatted according to the pre-established protocols for linkage disequilibrium (LD) score regression and genomic structural equation modelling (SEM) [1, 2].

Criteria for inclusion were GWAS studies that had a sample size of >5000 and only included unrelated European ancestry samples [3]. Because this analysis sought to measure shared genetic architecture between traits, we also ensured that summary statistics had not been adjusted for heritable covariates as this procedure can bias genetic effect estimates [4]. Furthermore, we did not include GWAS summary statistics that had been analysed using linear mixed modelling (LMM) methods for their association testing due to their potential to produce differential LD estimates compared to traditional regression methods [5, 6].

To minimise bias from rare or poorly imputed alleles, we carried out uniform quality control according to the genomic SEM protocol [2]. SNPs with a minor allele frequency (MAF) over 1% and an imputation (INFO) score above 0.9 were retained. As not all GWAS summary statistics supply this information, we filtered SNPs to Hapmap 3 SNPs with the major histocompatibility complex (MHC) region removed.

#### Sample and phenotype descriptions

Detailed descriptions of the individual cohorts that make up the non-Neale lab GWAS summary statistics that we used for our analysis can be found in the original GWAS publications [7-10]. Traits downloaded via the Neale lab (http://www.nealelab.is/uk-biobank) used UK Biobank data. All data we used only included individuals of European ancestry. Below we provide a brief overview for each of the traits used and how the phenotypes were measured.

#### Published GWAS studies

##### *Alzheimer's disease (Lambert et al., 2013)*

To model Alzheimer's disease (AD) we used the publicly available summary statistics for the stage 1 analysis from the GWAS performed by Lambert and colleagues [7]. The data comprised a meta-analysis of datasets from 4 large consortia: ADGC (10,273 cases and 10,892 controls), CHARGE (1,315 cases and 12,968 controls), EADI (2,243 cases and 6,017 controls) and GERAD (3,177 cases and 7,277 controls). This resulted in a final dataset consisting of 17,008 clinically diagnosed cases of AD and 37,154 controls (total sample size 54,162). The sample prevalence was therefore 31% and we used a population prevalence of 5% based on the age-specific prevalence of AD found in European populations aged over 50 years in a large meta-analysis study [11]. Association was analysed using logistic regression and included age, sex and principal components (PCs) as covariates.

##### *Major depressive disorder (Wray et al., 2018)*

We used the summary statistics from the GWAS performed by Wray et al [8] on major depressive disorder. The publicly available data included results of a meta-analysis of data from 6 large cohorts: PGC29 (16,823 cases and 25,632 controls), deCODE (1,980 cases and 9,536 controls), Generation Scotland (997 cases and 6,358 controls), GERA (7,162 cases and 38,307 controls), iPSYCH (18,629 cases and 17,841 controls) and UK Biobank (14,260 cases and 15,480 controls). Data used in the original study from 23andMe was excluded from this dataset so was not included in our analysis. This resulted in a sample size of 59,851 cases and 113,154 controls, forming an overall sample of 173,005 individuals. The sample prevalence was therefore 35% and we used a population prevalence of 13% taken from the Lancet Commission on Dementia Prevention, Intervention and Care [12]. Clinically diagnosed major depressive disorder was assessed by either a structured clinical review or extracted from electronic health records for all cohorts apart from the UK Biobank participants. For the UK Biobank participants, depression status was either taken from electronic health records or via self-report of major depression symptoms or treatment. Association was analysed using logistic regression, and principle component analysis was used to adjust for population stratification using the Psychiatric Genomics Consortium RICOPILI pipeline or similar methods.

***Insomnia (Jansen et al., 2019)***

We used the summary statistics for insomnia that were produced in the GWAS on sleep traits carried out by Jansen et al [9]. The publicly available data from this study only included the samples from UK Biobank and excluded 23andMe data. The trait for insomnia was characterised by the response to the question 'Do you have trouble falling asleep at night or do you wake up in the middle of the night?' The possible responses were 'never/rarely,' 'sometimes,' 'usually' or 'prefer not to answer.' For the GWAS analysis, cases were participants who answered 'usually' (109,402) and controls were those who answered 'never/rarely' or 'sometimes' (277,131), producing a total sample size of 386,533. This gave a sample prevalence of 28% and we based our population prevalence value of 32% on the prevalence of sleep disturbance in a Dutch cross-sectional population study of individuals aged 18-70 years [13]. GWAS association analysis was performed using logistic regression and was adjusted for age, sex, genotype array and 10 PCs.

***Type 2 diabetes mellitus (Scott et al., 2017)***

We used summary statistics from the Stage 1 GWAS analysis performed by Scott et al [10] for type 2 diabetes mellitus. This analysis was based on data from 18 studies within the DIAGRAM consortium and included 26,676 clinically diagnosed type 2 diabetes cases and 132,532 healthy controls. This resulted in a total sample size of 159,208 individuals and a sample prevalence of 17%. We based our population prevalence figure of 6% on the rate given in the Lancet Commission report [12]. The association analysis was performed using logistic regression and was adjusted for age, sex and ancestral PCs, but was not adjusted for BMI.

#### UK Biobank traits (all Neale lab, *http://www.nealelab.is/uk-biobank*)

The following traits are all based on data from UK Biobank, a UK-based cohort study of 500,000 participants from the general population aged 40-69 years at recruitment [14]. Inclusion of multiple traits measured within this cohort was appropriate as genomic SEM has previously been shown to be unbiased by sample overlap [2]. The GWASs were completed by the Neale lab using a standard pipeline developed by the research group and outlined in detail here (https://github.com/Nealelab/UK_Biobank_GWAS). In short, the GWASs were performed on data from unrelated participants of European ancestry and underwent uniform quality control procedures. Phenotypes were derived using the automated PHESANT pipeline [15]. We used the non-sex-stratified 'round 2' GWAS summary statistics that had been adjusted for the first 20 PCs, age, $age^2$, sex, age*sex and $age^2$*sex and were analysed using linear regression. As all traits were run on the same analysis pipeline to streamline such a high number of variables, the binary traits are therefore analysed as linear probability models, but for our analysis we calculated the appropriate sample and population prevalence to treat these traits as binary within our LD score regression and genomic SEM analyses. The appropriate sample prevalence figures for each binary trait was calculated using the sample size numbers provided within the Neale lab 'UKB SNP-Heritability Browser' (https://nealelab.github.io/UKBB_ldsc/h2_browser.html). Additional trait-specific characteristics are outlined under the subheadings below.

##### *Loneliness*

The loneliness phenotype is binary and based on participant response to the question 'Do you often feel lonely?' (UK Biobank Data Showcase code 2020). Those answering 'yes' were classified as cases (63,508) and those answering 'no' are controls (292,075). This resulted in a total sample size of 355,583 and a sample prevalence of 18%. We based our population prevalence estimate of 8% on the mean 10-year prevalence of loneliness published in a recent Age UK report based on data from the English Longitudinal Study of Ageing (ELSA) [16]. We felt this was a more reliable population estimate as it is longitudinal rather than cross-sectional and reflects a similar population. This phenotype was used as a proxy measure of social isolation for our analysis.

##### *Less social activity*

This binary phenotype was based on the Neale lab phenotype called 'Leisure/social activities: None of the above' and was based on the participant response to the question 'Which of the following do you attend once a week or more often?' (UK Biobank Data Showcase code 6160). The possible answers to this question were 'sports club or gym,' 'pub or social club,' 'religious group,' 'adult education class,' 'other group activity,' 'none of the above' or 'prefer not to answer.' We used the binary phenotype GWAS where the response 'None of the above' was classified as cases (108,704) and participants who selected one or more of the social or leisure activity responses were controls (251,359), giving an overall sample size of 360,063. Participants who responded with 'Prefer not answer' were designated as missing data and excluded from the sample in line with standard PHESANT procedures [15]. The sample prevalence was 30% and owing to the lack of a similar measure in external literature we also used this figure as the population prevalence.

As the questionnaire items were broad categories covering a wide range of social activities, we used this phenotype as a proxy for less social activity to model the practical aspect of social isolation in comparison to loneliness, which measures the emotional feeling of social isolation. We included both these traits to model the social isolation risk factor because previous literature has shown that these two aspects of social isolation may have differing effects on AD risk [17, 18].

***Hearing difficulty***

We used the binary phenotype called 'Hearing difficulty/problems with background noise' as our trait to model the risk factor hearing loss. This phenotype was based on the self-report question 'Do you find it difficult to follow a conversation if there is background noise (such as TV, radio, children playing)?' (UK Biobank Data Showcase code 2257). Participants who answered 'yes' were classified as cases (134,141) and those who answered 'no' were controls (219,842), giving an overall sample size of 353,983. Participants who had previously reported that they were completely deaf were not included in the sample. The sample prevalence was 38% and we used a comparable population prevalence estimate of 39%, which was based on the reported mean prevalence of self-reported hearing difficulty in participants aged over 50 in a UK-based prospective cohort study [19].

We used this phenotype instead of data for general hearing loss because difficulty in auditory scene analysis has been associated with AD so is potentially more relevant to dementia patients [20, 21]. Hearing difficulty with background noise also had a higher heritability estimate than the more general hearing problems phenotype within UK Biobank.

***Less education***

As we wanted to model the risk factor of having low educational attainment, we used the Neale lab GWAS phenotype named 'Qualifications: None of the above' as it dichotomised those who had educational qualifications versus those who had none. This trait was based on the participants' response to the question 'Which of the following qualifications do you have?' (UK Biobank Data Showcase code 6138). The possible answers were 'College or University degree,' 'A levels/AS levels or equivalent,' 'O levels/GCSEs or equivalent,' 'CSEs or equivalent,' 'NVQ or HND or HNC or equivalent,' 'Other professional qualifications e.g. nursing, teaching,' 'None of the above' or 'Prefer not to answer.' For the GWAS, these responses were converted into a binary phenotype where those answering 'None of the above' were classified as cases (61,093) and those answering any of the qualification types were controls (296,456), producing an overall sample size of 357,549. Participants who answered 'Prefer not to answer' were excluded from the analysis within the PHESANT pipeline [15]. The sample prevalence of no qualifications was 17%. As there are significant age-specific changes in educational attainment rates within the population due to legal and societal changes, we based our population prevalence on findings from a study that used data from the US-based Health and Retirement Study. We calculated the mean rate of having ≤11 years of education (i.e. did not complete high school) measured across two sets of individuals aged over 65, one which was assessed in 2000 and the latter completed in 2010, giving an average population prevalence of 27% [22].

***Physical inactivity***

To model the risk factor of physical inactivity we used the Neale lab GWAS titled 'Types of physical activity in last 4 weeks: None of the above.' This phenotype was based on the question 'In the last 4 weeks did you spend any time doing the following?' (UK Biobank Data Showcase code 6164). The possible responses were 'Walking for pleasure (not as a means of transport),' 'Other exercises (e.g. swimming, cycling, keep fit, bowling),' 'Strenuous sports,' 'Light DIY (e.g. pruning, watering the lawn),' 'Heavy DIY (e.g. weeding, lawn mowing, carpentry, digging),' 'None of the above' or 'Prefer not to answer.' Participants who answered 'Prefer not to answer' were excluded from the GWAS sample using PHESANT [15]. Individuals who answered 'None of the above' were classed as cases (21,255), whereas those who answered one or more of the types of physical activity were controls (338,008), producing an overall sample size of 359,263. Therefore, the sample prevalence of physical inactivity was 6%. We used a higher population prevalence estimate of 18% that was based on rates in the Lancet Commission report [12].

***Smoking***

This binary phenotype was named 'Smoking status: Current' and was based on the self-reported smoking status of the participants (UK Biobank Data Showcase code 20116). Cases were defined as individuals who were current smokers (37,088) and controls were individuals who had either never smoked or had smoked in the past (322,618), giving a sample size of 359,706. The sample prevalence was 10% and we used a population prevalence of 27% based on reported estimates in the Lancet Commission report [12]. Although the control group included past smokers, we felt that the members of this group were sufficiently different from current smokers as they had given up smoking at some stage and may not have smoked for a long period of their life. Furthermore, smoking during later life in particular has been associated with dementia so current smokers within this cohort are more representative of smoking as a dementia risk factor [12]. As the UK Biobank is a cohort study of participants aged above 40, we felt that those who had continued smoking displayed riskier health behaviours and were likely to have smoked for a long period, so including them as the case group for the smoking risk factor was preferable and likely to be a more homogenous phenotype.

***Alcohol intake frequency***

This ordinal phenotype was based on the participant response to the question 'About how often do you drink alcohol?' (UK Biobank Data Showcase code 1558). The possible responses were 'Daily or almost daily,' 'Three or four times a week,' 'Once or twice a week,' 'One to three times a month,' 'Special occasions only,' 'Never' or 'Prefer not to answer.' As with the other phenotypes, participants who answered 'Prefer not to answer' were excluded from the GWAS analysis. This ordinal phenotype was analysed using linear regression and analysed on the observed scale so did not require the sample or population prevalence to be specified. The total sample size was 360,726 individuals.

***Body mass index***

This continuous phenotype is based on the body mass index (BMI) measure that was completed by a study researcher during the initial assessment visit using an impedance device (UK Biobank Data Showcase code 23104). The phenotype is ordered by increments of 0.1 and was rank-normalised for GWAS analysis. The total sample size in the GWAS is 354,831 individuals and was analysed using linear regression and measured on the observed scale for our analysis. We included this trait as a proxy for the risk factor obesity.

***Deprivation status***

As a measure for deprivation status we used the GWAS summary statistics for the continuous phenotype of Townsend deprivation index at recruitment (UK Biobank Data Showcase code 189). The Townsend deprivation index was calculated using national census data and estimates the deprivation of each area by combining data on unemployment, non-car ownership, non-home ownership and household overcrowding [23]. Higher scores indicate higher levels of deprivation. Each UK Biobank participant was allocated a score that corresponded to their postcode at the time of recruitment. The phenotype was rank-normalised for GWAS and analysed using linear regression. The total sample size was 360,763 individuals.

***Systolic blood pressure***

We used the continuous phenotype that measured systolic blood pressure as our proxy for the risk factor of hypertension. The GWAS for this phenotype was based on the automated systolic blood pressure reading for each participant completed during their initial assessment visit (UK Biobank Data Showcase code 4080). The phenotype is continuous and has been rank-normalised for GWAS and analysed using linear regression. The total sample size is 340,159 individuals. We felt that using a continuous measure of systolic blood pressure was a more reliable measure than using data from a GWAS of clinically diagnosed hypertension because the diagnosis of hypertension can be subjective and not all individuals with hypertension receive a diagnosis, whereas all of the UK Biobank participants had their blood pressure measured during their baseline assessment. Therefore, it seemed likely that using a routine measure of blood pressure in a large population sample would provide more power to detect shared genetic architecture associated with increasing blood pressure levels.

#### Factor analysis

##### *Criteria to choose number of factors to extract for exploratory factor analysis*

We considered several parameters of the exploratory factor analysis (EFA) results to select how many factors we then specified in our follow-up confirmatory factor analysis (CFA). We set an *a priori* cut-off to include only factor loadings (i.e. the correlation coefficient between the latent factor and the observed trait) ≥0.20 when we specified the parameters of the subsequent CFA and we classed factor loadings ≥0.40 as highly stable [24, 25]. The predefined criteria to select the best model were that it should explain the most total genetic variance whilst ensuring that it only included factors that had 3 or more positive loadings ≥0.20 (including ≥2 'highly stable' loadings) and did not include any factor loadings >1.0 (i.e. a Heywood case).

##### *Assessing model fit for confirmatory factor analysis*

To assess the fit of the different models at CFA, we compared the results of several model fit tests in line with previously recommended cut-off points for genomic SEM [2]. This included the comparative fit index (CFI) test (with values ≥0.90 indicating moderate fit and values ≥0.95 indicating good fit) and the standardised root mean square residual (SRMR) (with values ≤0.10 indicating moderate fit and values below ≤0.05 indicating good fit) as our measures of absolute fit, and the Akaike information criterion (AIC) as our measure of relative fit (with lower values indicating better fit). We also compared $\chi^2$ statistics as a measure of exact fit (with lower values indicating better fit) [2].

***Structural equation modelling***

Our CFA models indicated that the 3 factors were moderately correlated, making it more difficult to interpret distinct clusters of potential shared genetic pathways between the included traits. We therefore tested several additional models to try to quantify the overarching common variation in a more meaningful way.

Firstly, to ascertain the level of common genetic variance between all our included traits we tested the performance of a common factor model (i.e. a model where there is only one latent factor of covariance across all measured traits). Next, we assessed the model fit for a second-order model (i.e. a model with a higher level latent factor that represents the common variance between the lower level latent factors) so that we could depict the inter-factor correlation as an additional higher level latent construct that encompasses the inter-factor correlation displayed in the CFA. Lastly, we tested the fit of a bi-factor model (i.e. a model where there is a latent factor of common variance across all traits as well as uncorrelated latent factors that represent covariance between sub-clusters of the measured traits). The benefit of a bi-factor model is that the factors are orthogonal (i.e. uncorrelated), since it accounts for the inter-factor correlation within the Common Factor as well as measuring additional latent constructs as clusters of distinct genetic covariance between specific measured traits. This was important in the current analysis since it made it easier to interpret the results because we could identify distinct clusters of shared genetic architecture between subgroups of our included traits, which enabled us to better theorise what they might each represent.

#### Post-hoc sensitivity analyses

##### *Chromosome-specific multivariable LD score regression*

Since the loadings for AD fluctuated substantially between EFA and CFA tested in the two halves of the autosomes, we ran post-hoc multivariable LD score regressions to measure the SNP-based heritability estimates and pairwise genetic correlations for the odd autosomes, even autosomes and all autosomes to assess whether this was due to substantial differences in effect across the chromosomal groups, which would lead to differences in correlation coefficients for factor analysis. We also calculated these estimates for each individual chromosome (chromosomes 1-22) to see how AD behaved compared to the more polygenic traits in our model.

##### *Exploratory factor analysis comparison across chromosomal groupings*

In view of the substantial differences in the heritability estimates and genetic correlation results between the odd and even chromosomal groups in the sensitivity LD score regression analysis, we ran additional EFA to compare the factor loading values for a 3-factor model based on different sets of chromosomes (odd vs. even vs. all).

##### *Genomic SEM of AD risk (excluding AD)*

As our sensitivity analysis revealed that the factor loadings for AD were only present in the odd autosomal data, we wanted to estimate the model with AD excluded. In addition, although the EFA loadings for the AD risk factors remained similar between odd and even chromosomes, there were some minor differences in the loadings that passed our cut-off value of ≥0.20 for inclusion into the CFA specification. Therefore, we ran an EFA for the 12 risk factors (AD and systolic blood pressure were excluded) using data from all of the autosomes and performed 3-factor CFA and SEM with the same cut-off properties as the main model to see how similar the performance of this model was compared to our initial one based on data from the odd autosomes with AD included.

### Supplementary Results

#### Exploratory factor analysis

We tested 2-, 3- and 4-factor models using EFA in the odd autosomes and found that the 3-factor model fitted the data best (**Tables S2-S4**). Although the 2-factor model had the most highly stable factor loadings (≥0.40), it explained less overall genetic variance compared to the 3-factor model (40.6% vs 42.5%) and captured less AD-specific variance (8.6% vs. 19%). The 4-factor model explained the most overall genetic variance (52.1%), but produced a factor loading of 1.088 for type 2 diabetes on the 4$^{th}$ factor. A loading value >1 is known as a Heywood case and represents an impossible value since it is not possible to explain over 100% of variance, so indicates that too many factors had been extracted for the 4-factor model [26, 27]. Therefore, we decided to base our subsequent CFA and SEM analyses on the output for the 3-factor model.

#### Sensitivity analysis results

##### *Chromosome-specific multivariable LD score regression*

The SNP-based heritability estimates for the odd autosomes, the even autosomes, all autosomes and individual chromosomes (1-22) are displayed in **Table S5**. All the traits except AD maintained high heritability Z scores (i.e. above 4) that did not differ substantially between the estimates calculated in the odd autosomes and the even autosomes. The heritability estimates were all <1% different between the odd and even autosomes, including for AD (except for physical inactivity, which had a difference of 1.13%). However, the heritability Z score for AD in the odd autosomes was only 2.23529 (which is below our cut-off of 4) compared to a Z score of 4.71014 in the even autosomes. This difference is due to the presence of a higher standard error for the odd autosomal estimate compared to the even autosomes (0.0102 vs. 0.0069).

In addition, the heritability estimates for the individual chromosomes fluctuated more for AD than for the other traits and displayed very low heritability Z scores (all <4 for AD). AD was also the only trait with negative heritability estimates for individual chromosomes (chromosomes 9, 13, 18 and 21).

The relative instability of the AD estimates compared to the other included traits are also reflected in the results of the pairwise genetic correlation estimates in different chromosomes. **Figure S3a** shows a heatmap of the pairwise genetic correlations measured in the odd autosomes compared to the estimates measured in the even autosomes only. Most traits show very similar estimates between both analyses and the correlation estimates have the same direction of effect between the odd and even autosomes. However, the pairwise correlation estimates between AD and its risk factors are substantially different between the odd and even autosomes. In many cases the two halves of the autosome have differential directions of correlation (the odd autosomes generally have positive correlations whereas more of the even autosomal correlations are negative correlations). **Figure S3b** shows a heatmap of the pairwise genetic correlations across all autosomes (i.e. an averaged effect between the odd and even correlations). Whereas the risk factor traits estimates are largely similar between the odd autosomes and all autosomes, the correlation estimates for AD are substantially lower in all autosomes compared with the odd autosomes. The magnitude of this difference is also reflected when measuring genetic correlations in individual chromosomes, with correlations between AD and risk factors changing dramatically by chromosome compared to genetic correlations between the risk factor traits (**Figure S4**).

***Post-hoc exploratory factor analysis***

EFA loadings for a 3-factor model in all 14 included traits showed that the loading estimates for most traits remained similar when measured in the odd autosomes, the even autosomes or all autosomes (**Table S6**). However, AD only showed factor loadings above our cut-off range (i.e. ≥0.20) in the odd autosomal EFA, whereas there were no loading estimates found for AD in the even autosomes and a subthreshold loading of 0.131 for Factor 1 when the EFA was run in all autosomes. Taken with our post-hoc LD score regression findings, these results indicate that it is inappropriate to split the chromosomes when performing genomic SEM with AD included, whereas it seems to be an appropriate method to guard against model overfitting if measuring the risk factor traits on their own. However, even though the loadings were similar for the AD risk factors, some of the loadings changed in relation to whether they met our EFA cut-off for inclusion in the CFA model.

***12-trait genomic SEM***

The 3-factor EFA of the 12 risk factors (AD and systolic blood pressure were excluded) conducted using the covariance matrix estimated in the data from all autosomes produced similar factor loadings to the initial 14-trait EFA and explained 50.8% of the total genetic variance (**Table S7**).

CFA and SEM of the 12-trait 3-factor model based on the parameters from the post-hoc EFA conducted across all autosomes produced comparable model fit statistics to our original 13-trait model based on odd autosomal data (see **Table 3** in the main text for model fit statistics). Though the factors were predominantly the same as the original model, there were fewer cross-loadings in the post-hoc model (Factor 1 dropped its loadings for insomnia and loneliness; Factor 2 dropped its loadings for BMI, depression and hearing difficulty; Factor 3 remained unchanged). However, the loading estimates were similar to those measured in the original model and the common factor model produced the same results despite exclusion of AD (see **Figure S5** for the relevant path diagrams).

These results indicate that although there are minimal differences between our original 13-trait model and our 12-trait model, our initial model appears to capture sufficiently stable latent constructs of covariance between the AD risk factors despite being calculated using the odd autosomes only. Furthermore, as the patterns of loadings remained similar and the model fit was comparable, our examination of how these constructs relate to the known literature remains valid.

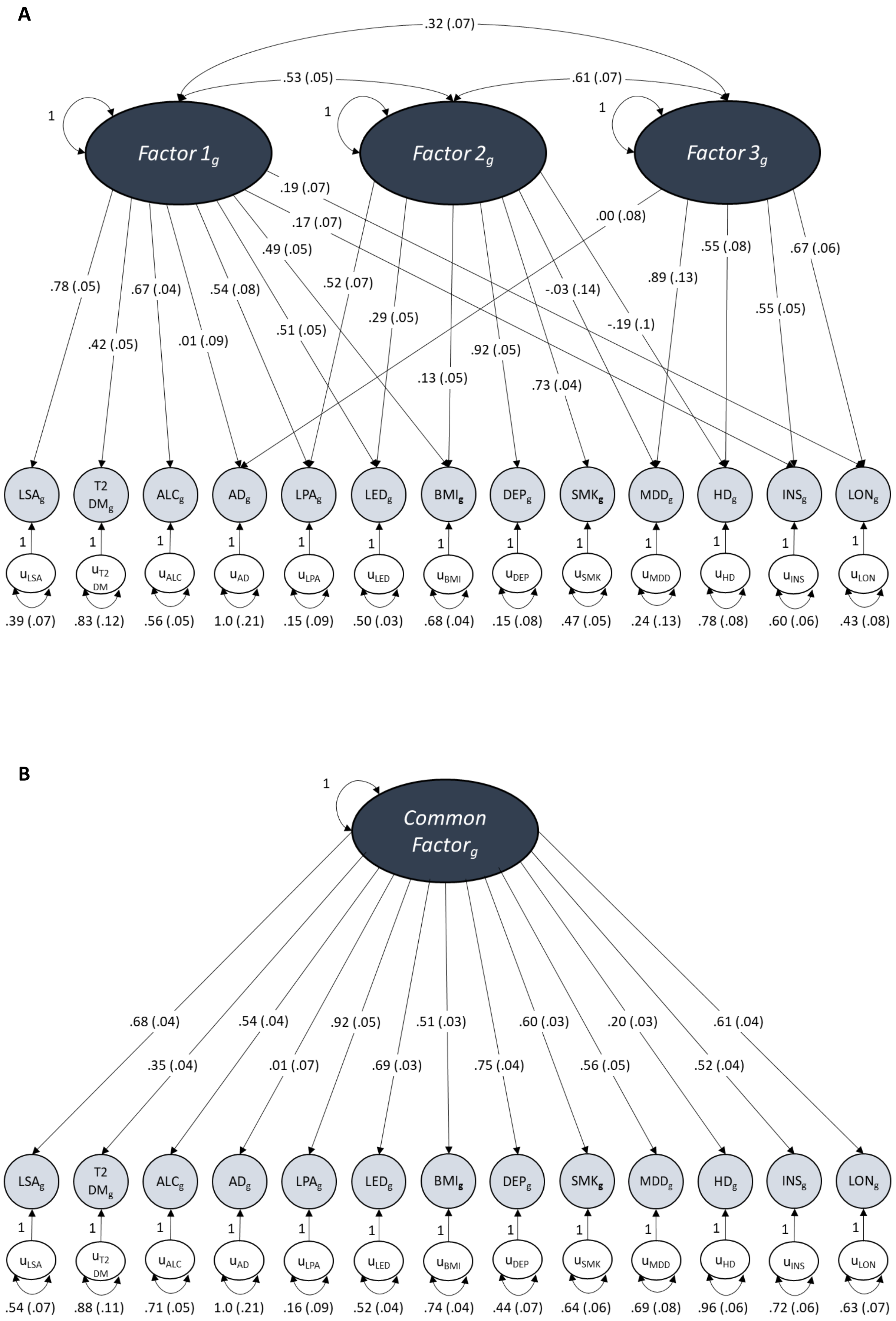
A
.32 (.07)
.53 (.05)
.61 (.07)
1
Factor 1g
Factor 2g
Factor 3g
.19 (.07)
.17 (.07)
.00 (.08)
.49 (.05)
.78 (.05)
.67 (.04)
.54 (.08)
.52 (.07)
.55 (.08)
.67 (.06)
.42 (.05)
.01 (.09)
.51 (.05)
.29 (.05)
-.03 (.14)
.89 (.13)
-.19 (.1)
.55 (.05)
.92 (.05)
.13 (.05)
.73 (.04)
LSAg
T2 DMg
ALCg
ADg
LPAg
LEDg
BMIg
DEPg
SMKg
MDDg
HDg
INSg
LONg
uLSA
uT2 DM
uALC
uAD
uLPA
uLED
uBMI
uDEP
uSMK
uMDD
uHD
uINS
uLON
.39 (.07)
.83 (.12)
.56 (.05)
1.0 (.21)
.15 (.09)
.50 (.03)
.68 (.04)
.15 (.08)
.47 (.05)
.24 (.13)
.78 (.08)
.60 (.06)
.43 (.08)
B
Common Factorg
.68 (.04)
.54 (.04)
.92 (.05)
.51 (.03)
.60 (.03)
.20 (.03)
.61 (.04)
.35 (.04)
.01 (.07)
.69 (.03)
.75 (.04)
.56 (.05)
.52 (.04)
.54 (.07)
.88 (.11)
.71 (.05)
1.0 (.21)
.16 (.09)
.52 (.04)
.74 (.04)
.44 (.07)
.64 (.06)
.69 (.08)
.96 (.06)
.72 (.06)
.63 (.07)

**C**

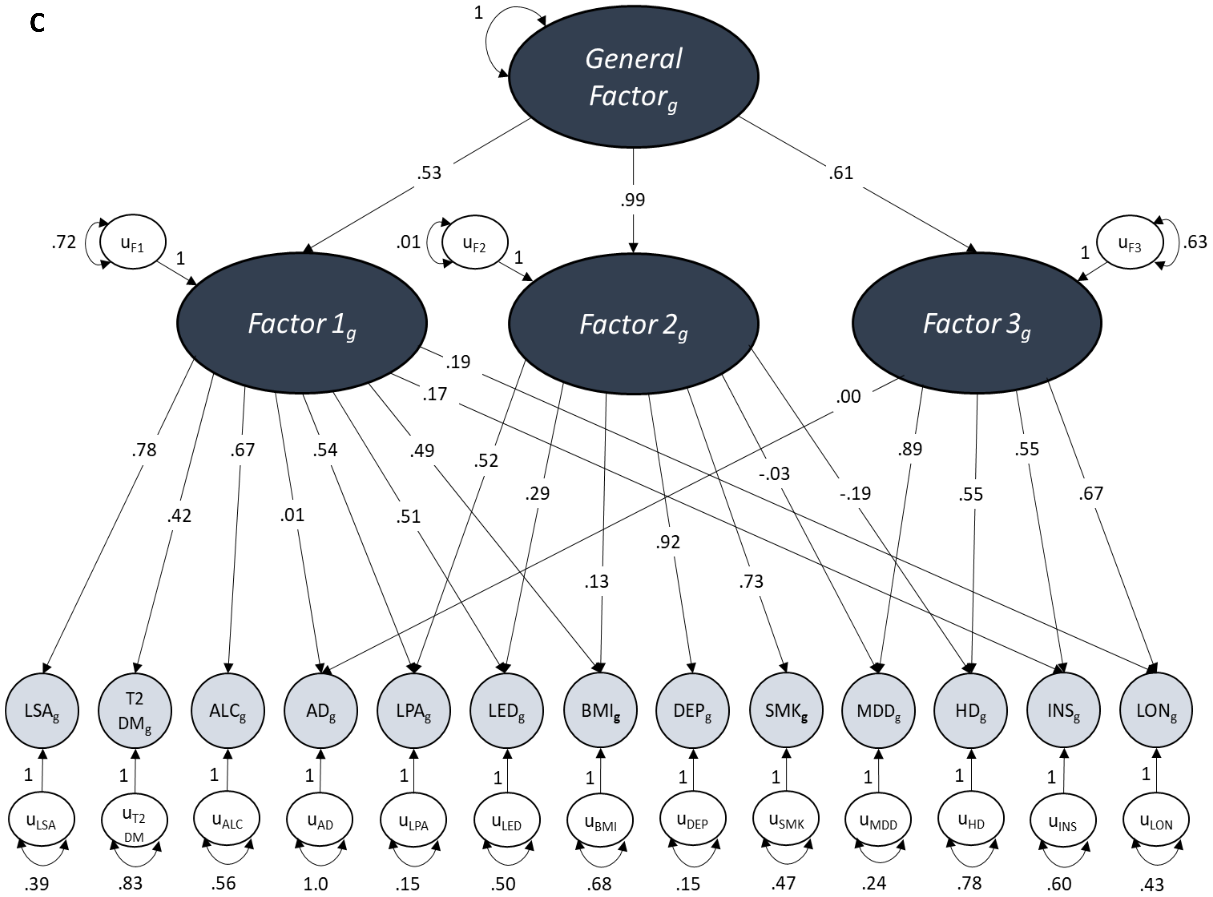


**D**

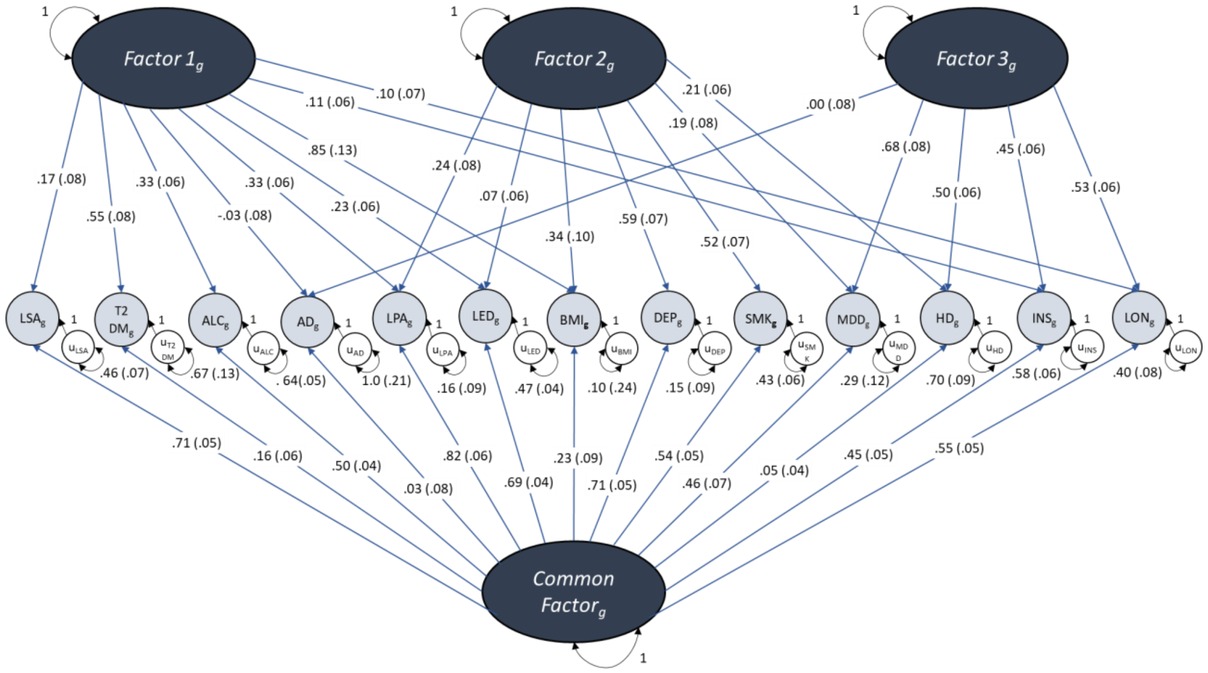


**Figure S1: Path diagrams of the standardised solutions for the (a) 3-factor CFA, (b) common factor model, (c) second-order model and (d) bi-factor model conducted across even autosomes only.**

AD Alzheimer’s disease; MDD major depressive disorder; INS insomnia; LON loneliness; LSA less social activity; HD hearing difficulty; LED less education; LPA physical inactivity; SMK smoking; ALC alcohol intake frequency; BMI body mass index; DEP deprivation status; T2DM type 2 diabetes mellitus.

A

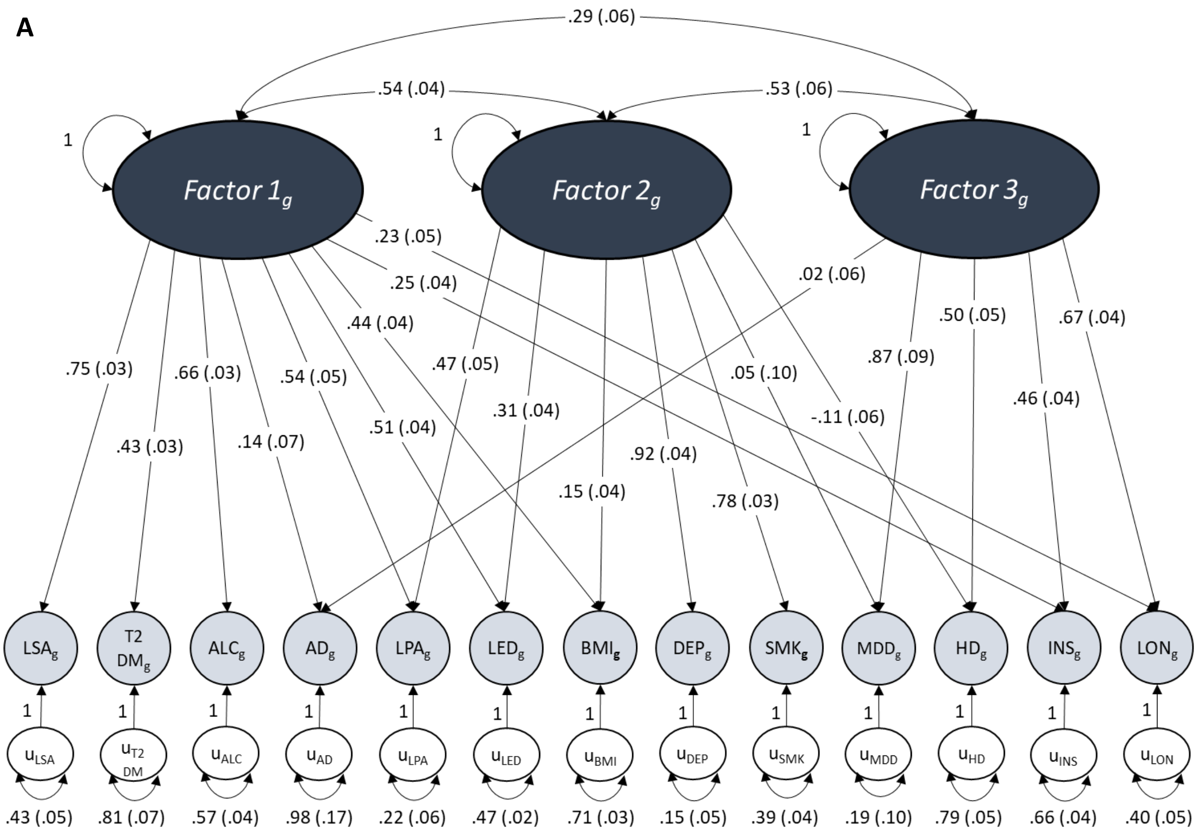

.29 (.06)
.54 (.04)
.53 (.06)
1
Factor 1g
1
Factor 2g
1
Factor 3g
.23 (.05)
.25 (.04)
.02 (.06)
.44 (.04)
.50 (.05)
.67 (.04)
.75 (.03)
.66 (.03)
.54 (.05)
.47 (.05)
.87 (.09)
.05 (.10)
.51 (.04)
.31 (.04)
.46 (.04)
.43 (.03)
.14 (.07)
-.11 (.06)
.92 (.04)
.15 (.04)
.78 (.03)
LSAg
T2 DMg
ALCg
ADg
LPAg
LEDg
BMIg
DEPg
SMKg
MDDg
HDg
INSg
LONg
uLSA
uT2 DM
uALC
uAD
uLPA
uLED
uBMI
uDEP
uSMK
uMDD
uHD
uINS
uLON
.43 (.05)
.81 (.07)
.57 (.04)
.98 (.17)
.22 (.06)
.47 (.02)
.71 (.03)
.15 (.05)
.39 (.04)
.19 (.10)
.79 (.05)
.66 (.04)
.40 (.05)


B

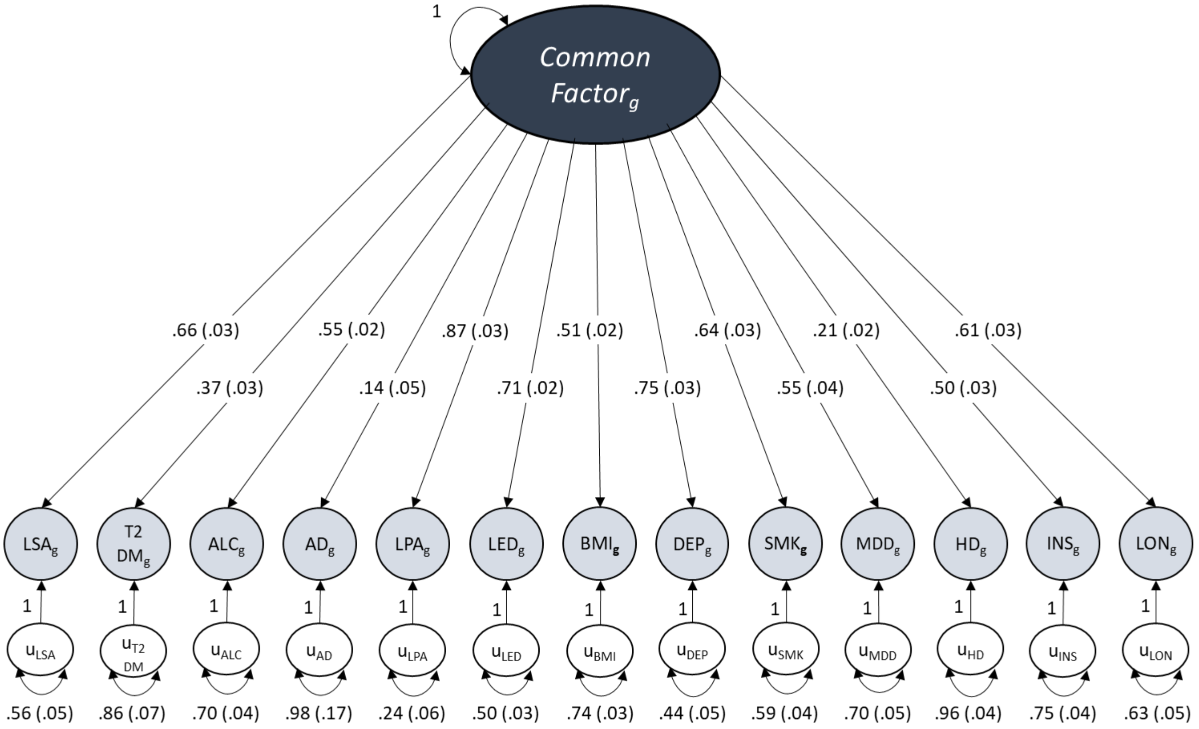

1
Common Factorg
.66 (.03)
.55 (.02)
.87 (.03)
.51 (.02)
.64 (.03)
.21 (.02)
.61 (.03)
.37 (.03)
.14 (.05)
.71 (.02)
.75 (.03)
.55 (.04)
.50 (.03)
LSAg
T2 DMg
ALCg
ADg
LPAg
LEDg
BMIg
DEPg
SMKg
MDDg
HDg
INSg
LONg
uLSA
uT2 DM
uALC
uAD
uLPA
uLED
uBMI
uDEP
uSMK
uMDD
uHD
uINS
uLON
.56 (.05)
.86 (.07)
.70 (.04)
.98 (.17)
.24 (.06)
.50 (.03)
.74 (.03)
.44 (.05)
.59 (.04)
.70 (.05)
.96 (.04)
.75 (.04)
.63 (.05)

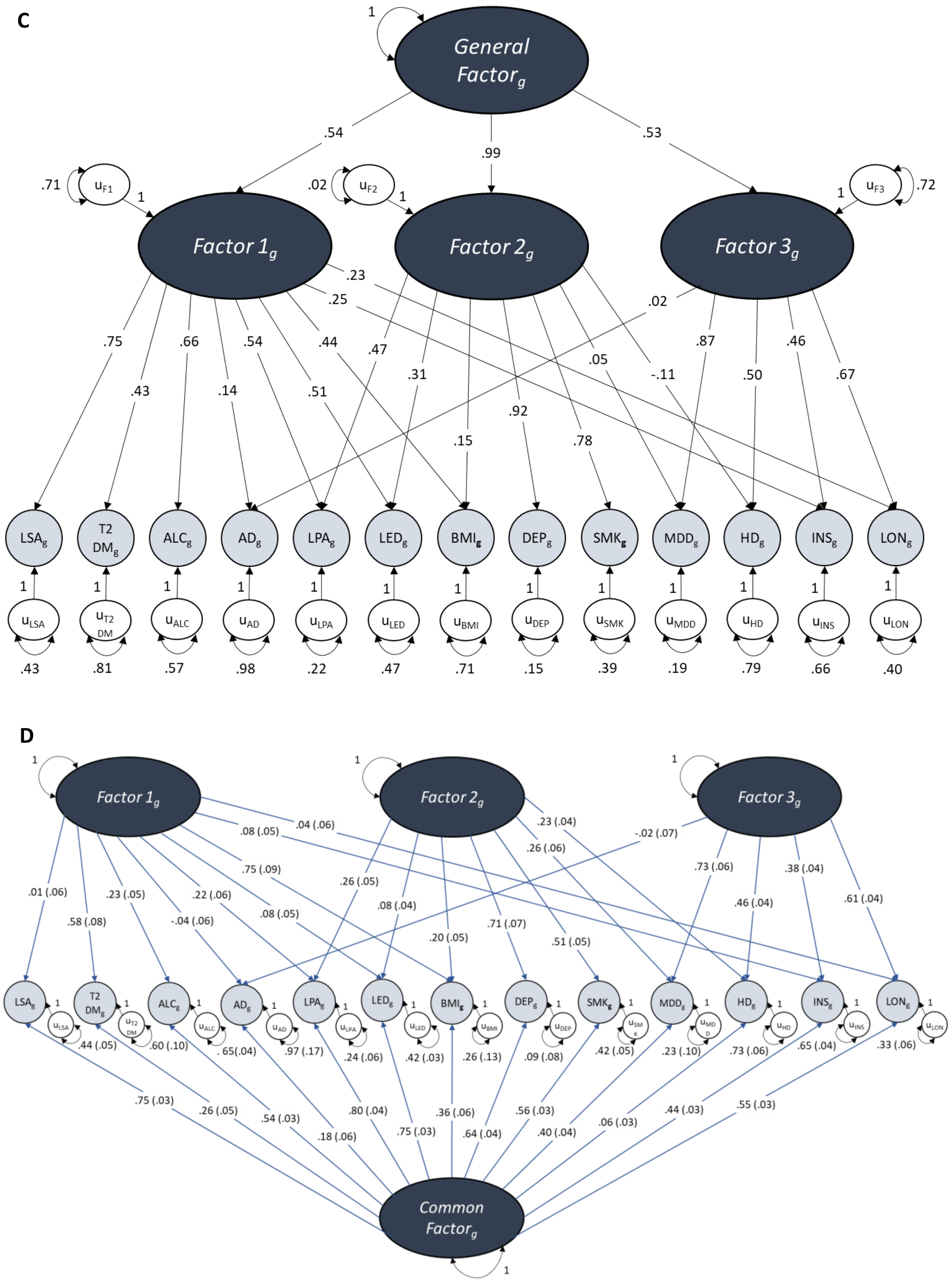


**Figure S2: Path diagrams of the standardised solutions for the (a) 3-factor CFA, (b) common factor model, (c) second-order model and (d) bi-factor model conducted across all autosomes except for chromosome 19.**

AD Alzheimer's disease; MDD major depressive disorder; INS insomnia; LON loneliness; LSA less social activity; HD hearing difficulty; LED less education; LPA physical inactivity; SMK smoking; ALC alcohol intake frequency; BMI body mass index; DEP deprivation status; T2DM type 2 diabetes mellitus.

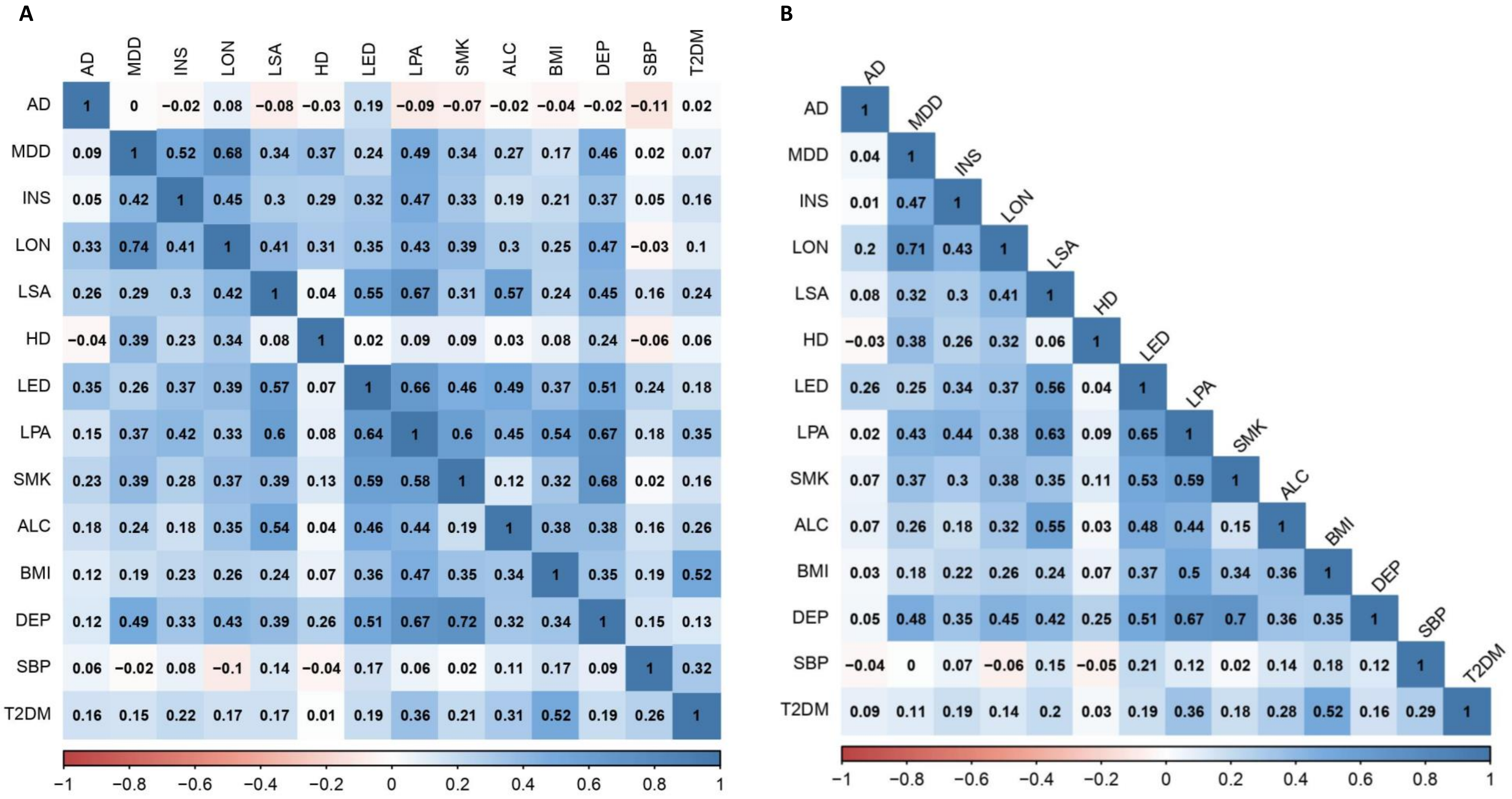


**Figure S3: Heatmaps comparing the pairwise genetic correlations measured in the odd and even autosomes (A) and all autosomes (B) using multivariable LD score regression.** The lower triangle of the matrix in Figure A represents the pairwise genetic correlations in the odd autosomes whereas the upper triangle represents the genetic correlations of the even autosomes. Figure B shows the pairwise genetic correlations across all autosomes (i.e. chromosomes 1-22).

AD Alzheimer's disease; MDD major depressive disorder; INS insomnia; LON loneliness; LSA less social activity; HD hearing difficulty; LED less education; LPA physical inactivity; SMK smoking; ALC alcohol intake frequency; BMI body mass index; DEP deprivation status; SBP systolic blood pressure; T2DM type 2 diabetes mellitus.

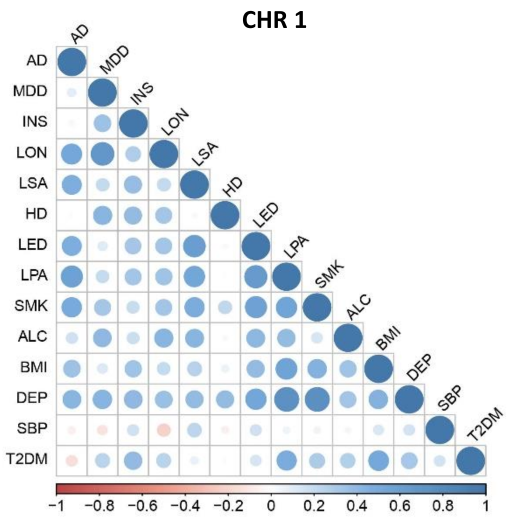
CHR 1
AD
MDD
INS
LON
LSA
HD
LED
LPA
SMK
ALC
BMI
DEP
SBP
T2DM
−1 −0.8 −0.6 −0.4 −0.2 0 0.2 0.4 0.6 0.8 1

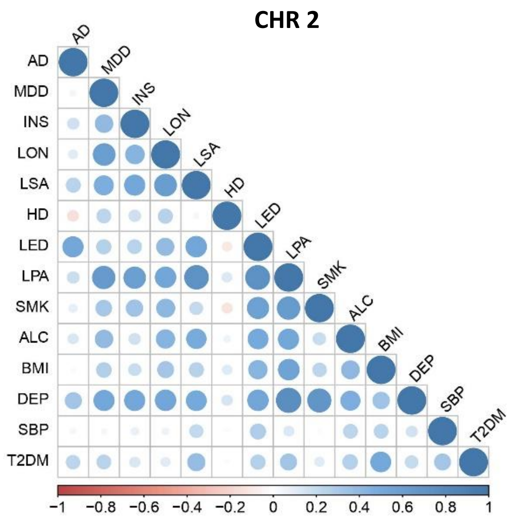
CHR 2
AD
MDD
INS
LON
LSA
HD
LED
LPA
SMK
ALC
BMI
DEP
SBP
T2DM
−1 −0.8 −0.6 −0.4 −0.2 0 0.2 0.4 0.6 0.8 1

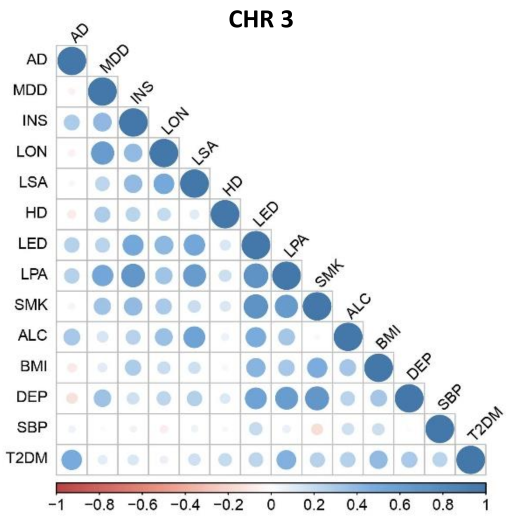
CHR 3
AD
MDD
INS
LON
LSA
HD
LED
LPA
SMK
ALC
BMI
DEP
SBP
T2DM
−1 −0.8 −0.6 −0.4 −0.2 0 0.2 0.4 0.6 0.8 1

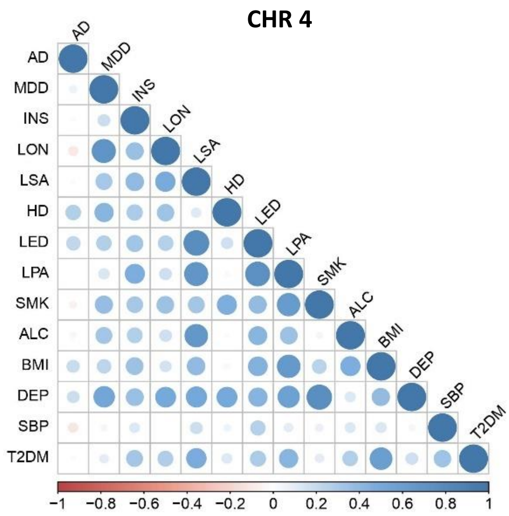
CHR 4
AD
MDD
INS
LON
LSA
HD
LED
LPA
SMK
ALC
BMI
DEP
SBP
T2DM
−1 −0.8 −0.6 −0.4 −0.2 0 0.2 0.4 0.6 0.8 1

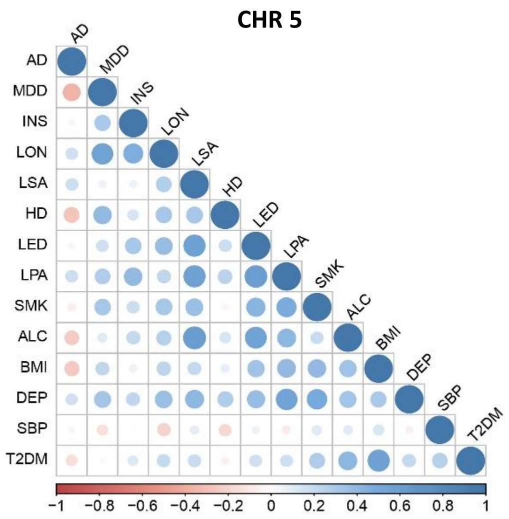
CHR 5
AD
MDD
INS
LON
LSA
HD
LED
LPA
SMK
ALC
BMI
DEP
SBP
T2DM
−1 −0.8 −0.6 −0.4 −0.2 0 0.2 0.4 0.6 0.8 1

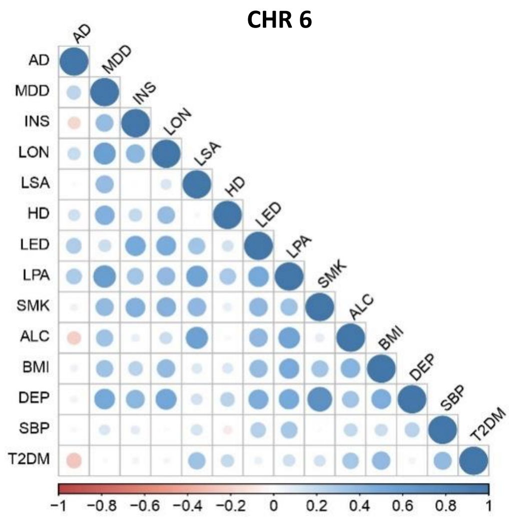
CHR 6
AD
MDD
INS
LON
LSA
HD
LED
LPA
SMK
ALC
BMI
DEP
SBP
T2DM
−1 −0.8 −0.6 −0.4 −0.2 0 0.2 0.4 0.6 0.8 1

**CHR 7**

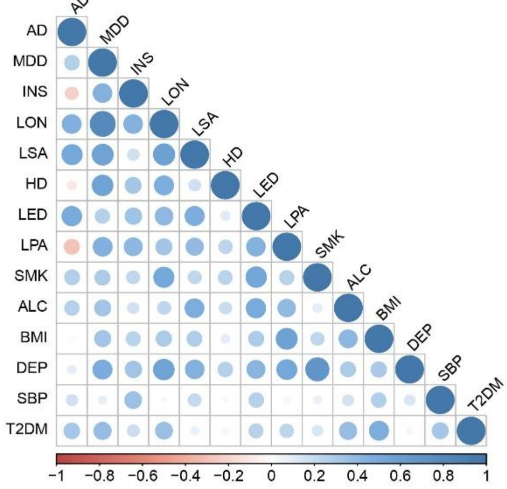


**CHR 8**

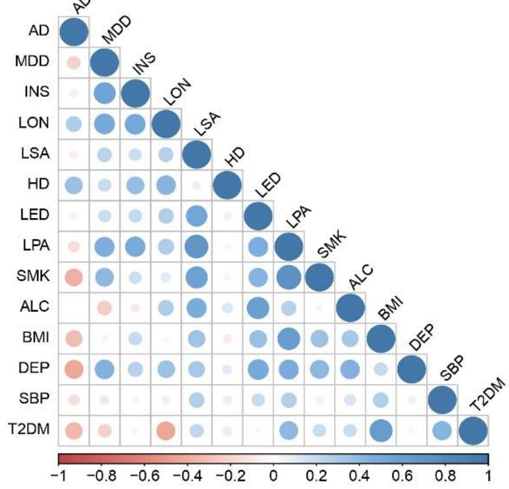


**CHR 9**

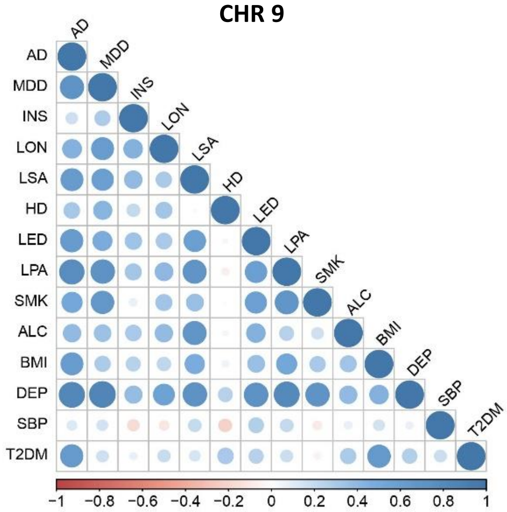


**CHR 10**

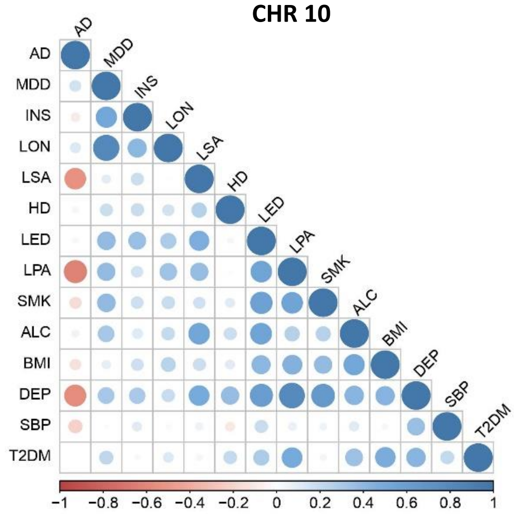


**CHR 11**

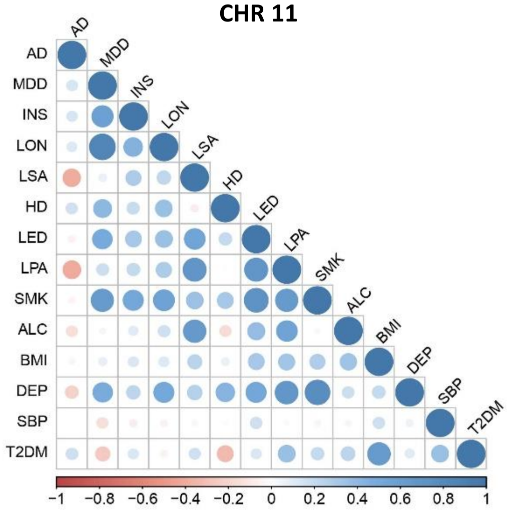


**CHR 12**

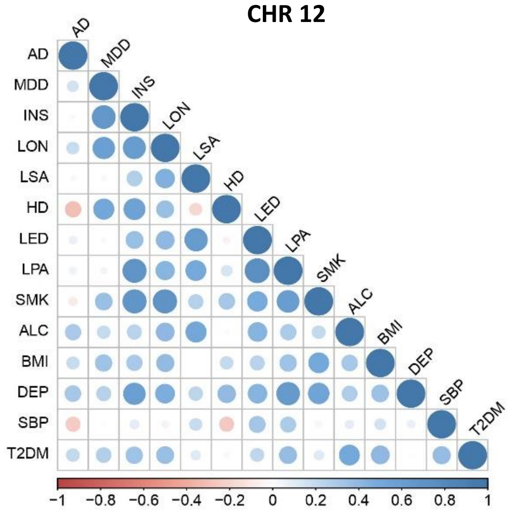

**CHR 13**

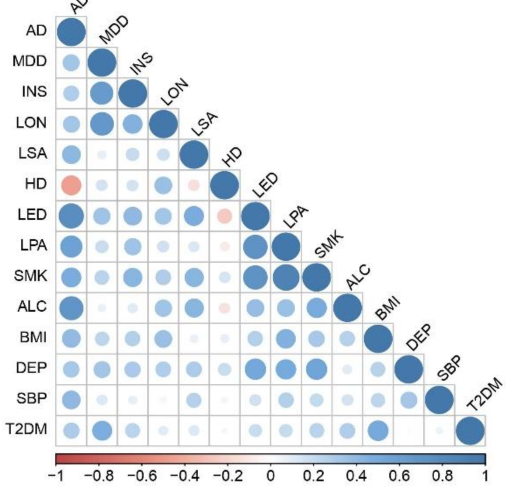


**CHR 14**

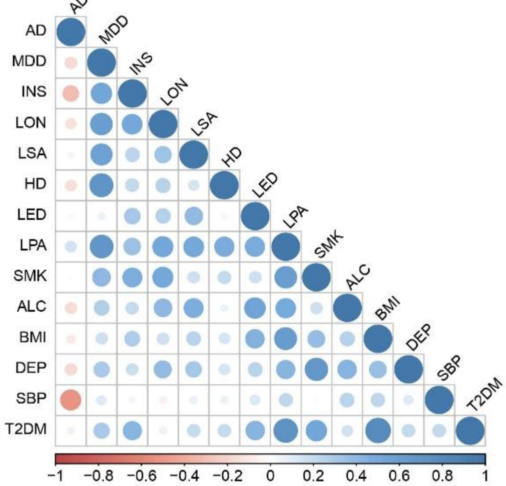


**CHR 15**

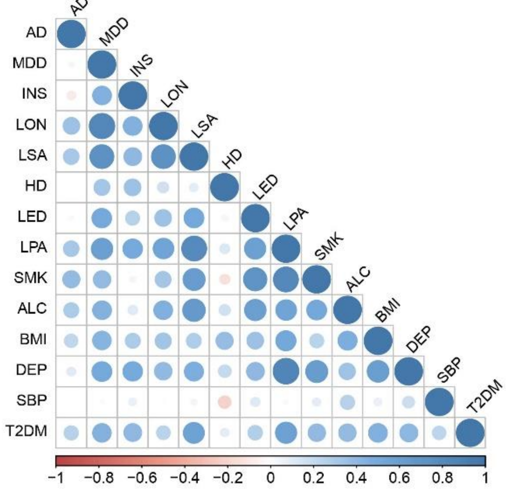


**CHR 16**

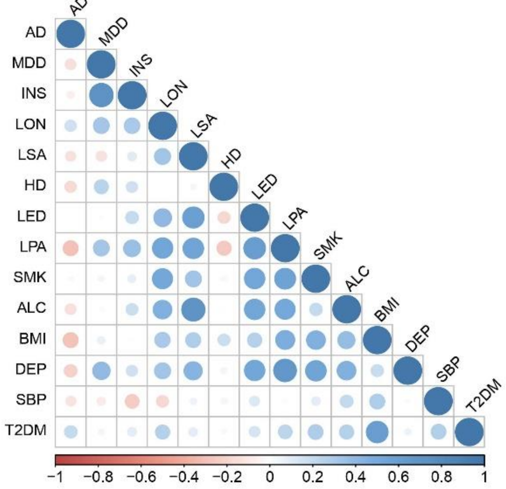


**CHR 17**

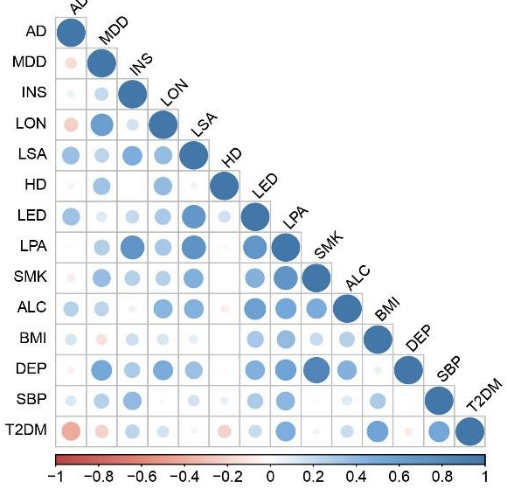


**CHR 18**

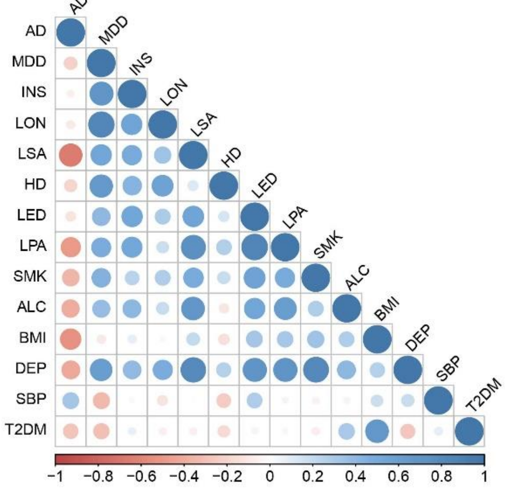

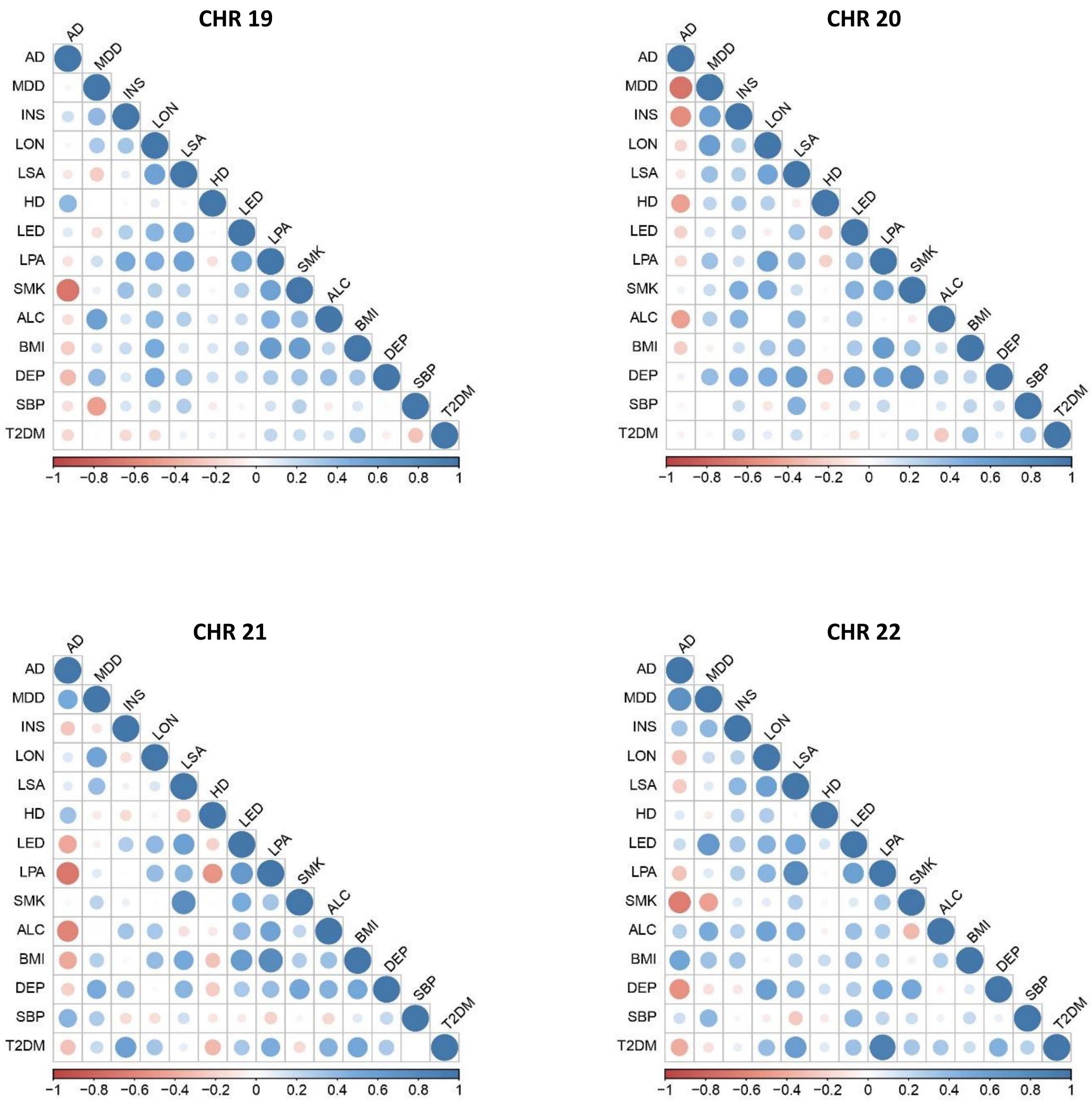


**Figure S4: Heatmaps of pairwise correlations across individual chromosomes using multivariable LD score regression.** Each matrix represents the pairwise genetic correlations for an individual chromosome (CHR), and the strength of correlation is signified by the size and shade of each circle. The stronger the correlation, the larger the circle and the darker the colour. Blue circles represent positive correlations and red circles denote negative correlations.

AD Alzheimer's disease; MDD major depressive disorder; INS insomnia; LON loneliness; LSA less social activity; HD hearing difficulty; LED less education; LPA physical inactivity; SMK smoking; ALC alcohol intake frequency; BMI body mass index; DEP deprivation status; SBP systolic blood pressure; T2DM type 2 diabetes mellitus.

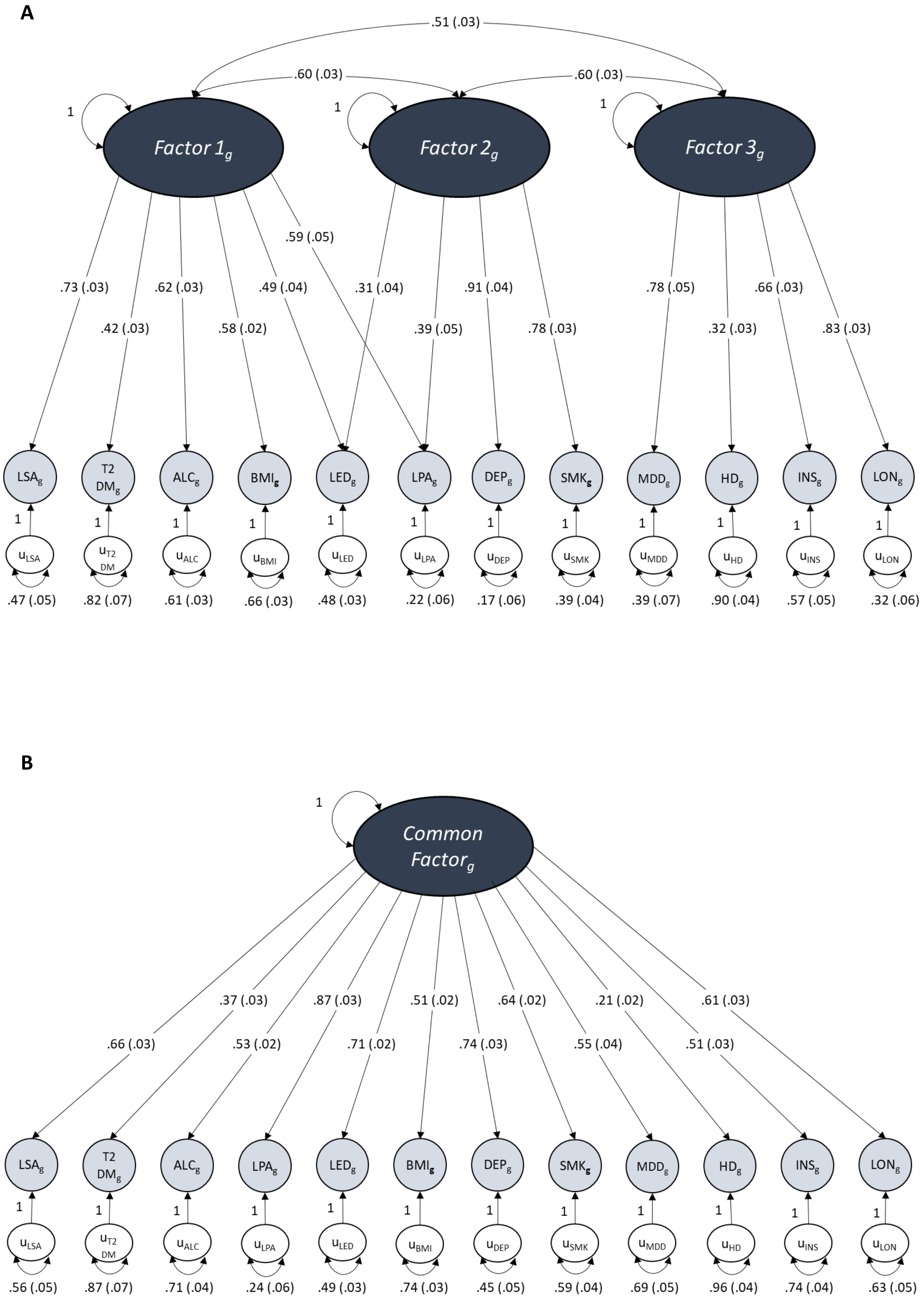
A
.51 (.03)
.60 (.03)
.60 (.03)
1
Factor 1g
Factor 2g
Factor 3g
.73 (.03)
.42 (.03)
.62 (.03)
.58 (.02)
.49 (.04)
.59 (.05)
.31 (.04)
.39 (.05)
.91 (.04)
.78 (.03)
.78 (.05)
.32 (.03)
.66 (.03)
.83 (.03)
LSAg
T2 DMg
ALCg
BMIg
LEDg
LPAg
DEPg
SMKg
MDDg
HDg
INSg
LONg
uLSA
uT2 DM
uALC
uBMI
uLED
uLPA
uDEP
uSMK
uMDD
uHD
uINS
uLON
.47 (.05)
.82 (.07)
.61 (.03)
.66 (.03)
.48 (.03)
.22 (.06)
.17 (.06)
.39 (.04)
.39 (.07)
.90 (.04)
.57 (.05)
.32 (.06)
B
Common Factorg
.66 (.03)
.37 (.03)
.53 (.02)
.87 (.03)
.71 (.02)
.51 (.02)
.74 (.03)
.64 (.02)
.55 (.04)
.21 (.02)
.51 (.03)
.61 (.03)
LSAg
T2 DMg
ALCg
LPAg
LEDg
BMIg
DEPg
SMKg
MDDg
HDg
INSg
LONg
uLSA
uT2 DM
uALC
uLPA
uLED
uBMI
uDEP
uSMK
uMDD
uHD
uINS
uLON
.56 (.05)
.87 (.07)
.71 (.04)
.24 (.06)
.49 (.03)
.74 (.03)
.45 (.05)
.59 (.04)
.69 (.05)
.96 (.04)
.74 (.04)
.63 (.05)

C

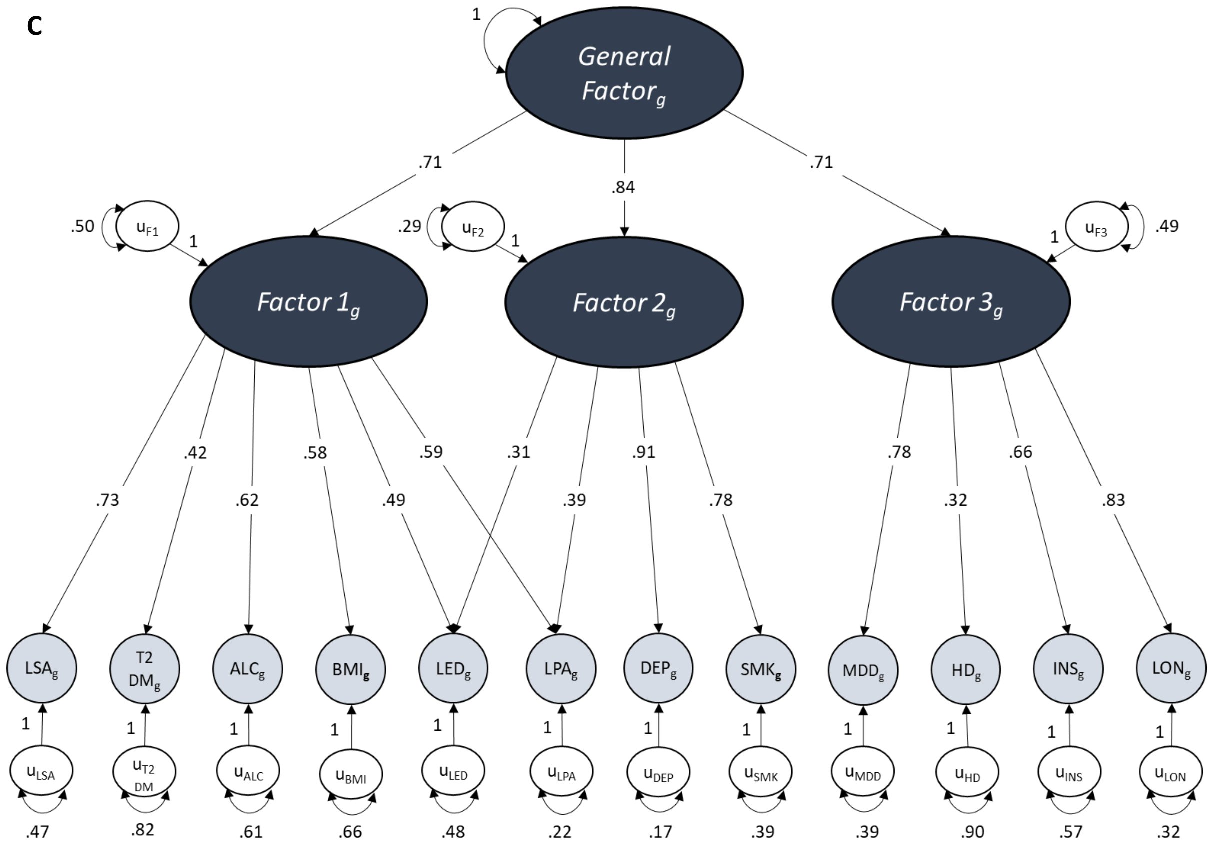
General Factorg
1
.71
.84
.71
Factor 1g
Factor 2g
Factor 3g
uF1
uF2
uF3
.50
.29
.49
1
1
1
.73
.42
.62
.58
.49
.59
.31
.39
.91
.78
.78
.32
.66
.83
LSAg
T2 DMg
ALCg
BMIg
LEDg
LPAg
DEPg
SMKg
MDDg
HDg
INSg
LONg
uLSA
uT2 DM
uALC
uBMI
uLED
uLPA
uDEP
uSMK
uMDD
uHD
uINS
uLON
.47
.82
.61
.66
.48
.22
.17
.39
.39
.90
.57
.32

D

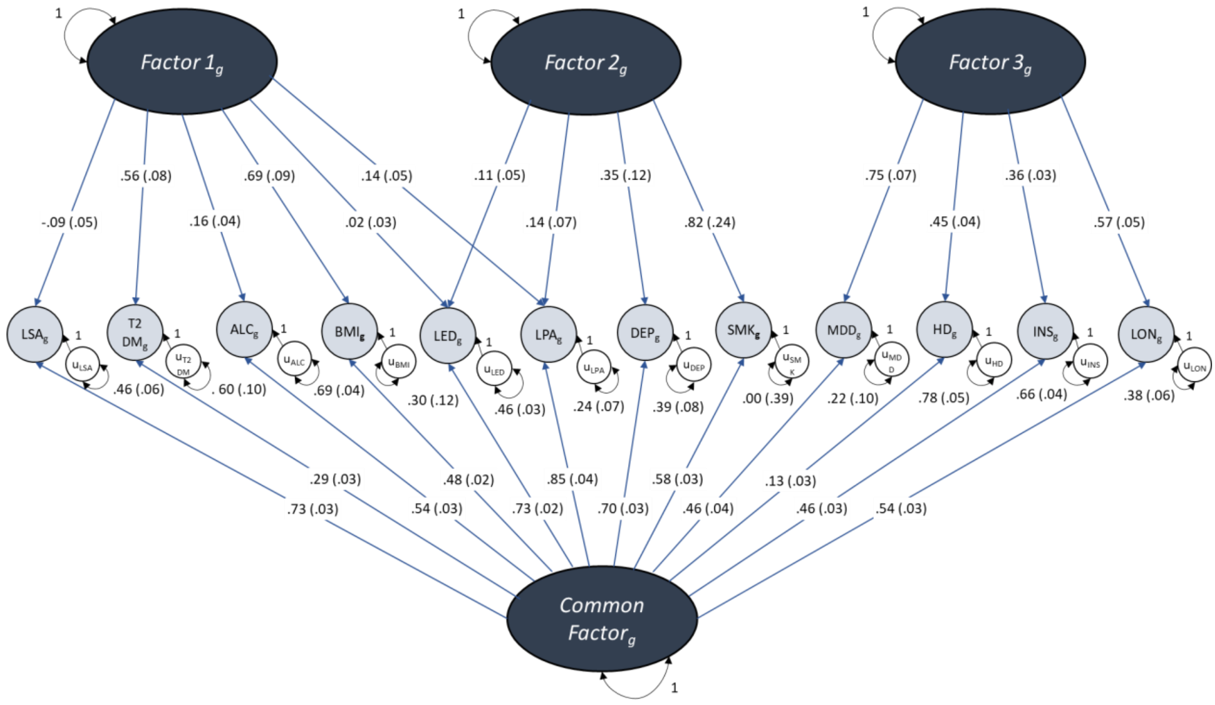
Factor 1g
Factor 2g
Factor 3g
1
1
1
-.09 (.05)
.56 (.08)
.16 (.04)
.69 (.09)
.02 (.03)
.14 (.05)
.11 (.05)
.14 (.07)
.35 (.12)
.82 (.24)
.75 (.07)
.45 (.04)
.36 (.03)
.57 (.05)
LSAg
T2 DMg
ALCg
BMIg
LEDg
LPAg
DEPg
SMKg
MDDg
HDg
INSg
LONg
uLSA
uT2 DM
uALC
uBMI
uLED
uLPA
uDEP
uSMK
uMDD
uHD
uINS
uLON
.46 (.06)
. 60 (.10)
.69 (.04)
.30 (.12)
.46 (.03)
.24 (.07)
.39 (.08)
.00 (.39)
.22 (.10)
.78 (.05)
.66 (.04)
.38 (.06)
.29 (.03)
.73 (.03)
.54 (.03)
.48 (.02)
.73 (.02)
.85 (.04)
.70 (.03)
.58 (.03)
.46 (.04)
.13 (.03)
.46 (.03)
.54 (.03)
Common Factorg
1

**Figure S5: Path diagrams of the standardised solutions for the post-hoc 12-trait (a) 3-factor CFA, (b) common factor model, (c) second-order model and (d) bi-factor model conducted across all autosomes.** The parameters for these models were based on a post-hoc EFA analysis calculated using data from all autosomes but with Alzheimer's disease and systolic blood pressure removed.

MDD major depressive disorder; INS insomnia; LON loneliness; LSA less social activity; HD hearing difficulty; LED less education; LPA physical inactivity; SMK smoking; ALC alcohol intake frequency; BMI body mass index; DEP deprivation status; T2DM type 2 diabetes mellitus.
